# Comparative effectiveness of gabapentin and pregabalin on reduction in alcohol use: A nationwide observational cohort study

**DOI:** 10.64898/2026.01.13.26344031

**Authors:** Tommy Gunawan, Joshua C. Gray, Mingjian Shi, Thomas Wingo, Aliza P. Wingo, David A. Fiellin, John Tazare, Mehdi Farokhnia, Lorenzo Leggio, Henry R. Kranzler, Christopher T. Rentsch

## Abstract

**Background:** Gabapentin and pregabalin have potential utility for treating alcohol use disorder (AUD), but their comparative effectiveness in reducing alcohol consumption in real-world settings is unknown. We compared changes in alcohol consumption associated with gabapentin or pregabalin treatment with those of matched unexposed comparators.

**Methods:** We identified patients who were prescribed gabapentin or pregabalin for ≥60 days for any indication between 1 January 2009 and 30 June 2022 using electronic health records from the Veterans Aging Cohort Study (VACS-National). Alcohol consumption was measured using routinely-collected Alcohol Use Disorders Identification Test-Consumption (AUDIT-C) questionnaires. We used propensity score matching to balance baseline characteristics between groups. Three comparisons were conducted: gabapentin vs. unexposed, pregabalin vs. unexposed, and pregabalin vs. gabapentin. Changes in AUDIT-C scores were estimated using multivariable difference-in-difference linear regression models. Analyses were stratified by baseline AUD diagnosis and AUDIT-C categories (lower-risk, at-risk, hazardous/binge).

**Results:** We identified 592,957 gabapentin initiators, 14,923 pregabalin initiators, and 2,959,006 eligible unexposed comparators who were eligible for matching. Compared to unexposed individuals, patients who received gabapentin (DiD: 0.09, 95% CI 0.06, 0.11, p<0.0001) or pregabalin (DiD: 0.14, 95% CI 0.02, 0.26, p=0.0279) reported greater reductions in AUDIT-C scores. When compared head-to-head, pregabalin initiators reported greater reductions in AUDIT-C scores than gabapentin initiators with the largest difference observed among those with AUD (DiD: 0.86, 95% CI 1.22, 0.50, p<0.0001) and those who report baseline hazardous/binge drinking (DiD: 1.74, 95% CI 2.21, 1.27, p<0.0001).

**Discussion:** In this large, nationwide cohort, treatment with gabapentin and pregabalin were associated with reductions in reported alcohol consumption, compared to matched unexposed comparators. Initiation of pregabalin was associated with greater reductions than with gabapentin, particularly among those with AUD and those with highest severity of alcohol use. Known safety concerns and risk of misuse should be considered when prescribing these medications. Randomized clinical trials directly comparing these medications are necessary to validate these findings.

## Introduction

Globally, alcohol use disorder (AUD) affects 400 million individuals, with 2.6 million deaths attributable to alcohol consumption ^1^. Over 28 million U.S. adults had past-year AUD in 2023, and excessive alcohol consumption costs approximately $250 billion in 2010 ^2,3^. In addition to behavioral treatments ^4^, three medications – disulfiram, acamprosate, and naltrexone - have been approved by the U.S. Food and Drug Administration for the treatment of AUD ^5^, though, their number and effectiveness are suboptimal ^6–8^. Drug repurposing is a potential strategy to address the need for novel AUD pharmacotherapies ^9–13^.

Recently, our group developed a genetically informed drug-repurposing pipeline that integrates genome-wide association study (GWAS) data with network-based methods and pharmacologic resources to identify drug repurposing candidates for AUD ^14^. Using the pipeline, we identified calcium channel subunit genes *CACNA1C*, *CACNA1D*, *CACNB2* and *CACNB3* as alcohol-related molecular targets. A recent study integrated brain proteogenomic data with AUD GWAS data identified *CACNA2D1* as a candidate causal protein for AUD ^15^. Together, these findings converge on calcium channel signaling as an underlying mechanism driving alcohol use risk and suggests that drugs targeting these channels may hold therapeutic potential for AUD. Gabapentin and pregabalin represent two drugs targeting these channels and for which preexisting literature suggests a potential pharmacotherapeutic role for AUD.

Gabapentin, a gabapentinoid approved for postherpetic neuralgia, partial seizures, and restless leg syndrome ^16–18^, has been investigated for AUD and used off-label for the treatment for alcohol withdrawal syndrome ^19^ and to reduce drinking in individuals with AUD ^20^. Gabapentin binds to the α_2_δ subunit of voltage-gated calcium channels ^21^. Some randomized clinical trials (RCTs) of gabapentin for treating AUD showed evidence that the drug reduced alcohol craving, number of drinking days and heavy drinking days, and increasing abstinence rates relative to placebo ^22–28^. However, other studies, including a large multi-site RCT that tested an extended-release formulation of gabapentin ^29^, failed to show an effect ^30^. A meta-analysis of seven RCTs comparing gabapentin to placebo showed a medium effect size of gabapentin reducing the percentage of heavy drinking days (Hedge’s g = −0.64), with smaller, non-significant effects on abstinence, relapse to heavy drinking and drinks per day ^20^. The American Psychiatric Association also recommends gabapentin (as well as topiramate) as second-line medication for AUD ^5^.

Pregabalin, also a gabapentinoid active at the α_2_δ subunit of voltage-gated calcium channels ^21^, is approved for the treatment of neuropathic pain, partial onset seizures, and fibromyalgia ^31–34^. Three open-label trials evaluating the efficacy of pregabalin for alcohol dependence or AUD showed that patients treated with pregabalin experienced a reduction in alcohol craving, withdrawal symptoms, and heavy drinking days ^35–37^. A single-blind RCT also showed that individuals prescribed pregabalin had higher rates of abstinence compared to those prescribed tiapride or lorazepam ^38^. However, evidence from double-blind RCTs is mixed. Pregabalin showed no effect on alcohol withdrawal symptoms relative to placebo ^39^, but a longer period of abstinence than naltrexone-treated individuals ^40^. The small study samples and inconsistencies in study design preclude drawing conclusions regarding the efficacy of pregabalin for AUD.

Despite similarities in the pharmacodynamic effects of gabapentin and pregabalin, there are notable differences in their pharmacokinetics and clinical use. Pregabalin has higher bioavailability and more predictable linear absorption than gabapentin, whose bioavailability decreases at higher doses ^21,41^. Thus, pregabalin achieves more consistent plasma concentrations and may require a lower dosage to achieve therapeutic efficacy. Pregabalin also has a faster onset of action, which may be advantageous for managing acute symptoms of alcohol withdrawal or craving ^21,41^. However, pregabalin carries a higher potential for abuse liability, misuse, and dependence, and is classified as a Schedule V controlled substance in the United States, while gabapentin is unscheduled ^42^, though this differs across states and countries ^43,44^. Regulatory differences impact prescribing practices and access, and gabapentin is less expensive and more widely available than pregabalin, whose controlled status may limit its availability despite a more favorable pharmacokinetic profile. Whether differences between gabapentin and pregabalin translate to differential effectiveness to treat AUD is unknown, as no study has directly compared their association with a reduction in alcohol use.

Thus, we investigated associations between gabapentin or pregabalin prescriptions for any indication and changes in alcohol use using data from the US Department of Veterans Affairs’ (VA) electronic health record (EHR). We evaluated whether associations differed by AUD diagnosis, AUDIT-C risk categories at baseline, and medication dosage. We hypothesized that receipt of gabapentin and pregabalin would each be associated with a reduction in alcohol use, and that this effect would be greater among individuals with AUD, those in higher AUDIT-C risk categories, and those who were prescribed higher doses of the medications. Given pregabalin’s greater bioavailability, faster onset of action, and more predictable pharmacokinetics than gabapentin’s, we also hypothesized that receipt of pregabalin would be associated with greater reductions in alcohol use than gabapentin in a head-to-head comparison.

## Methods

### Study Sample

We conducted an observational cohort study using data from the Veterans Aging Cohort Study (VACS-National), which includes all >14.1 million individuals who have received any VA care since 1999. The VA is the largest integrated U.S. healthcare system serving approximately 6 million patients annually in over 1,300 inpatient and outpatient medical centers. All care is recorded in the EHR with daily uploads to the VA Corporate Data Warehouse. Available data include demographics, International Classification of Diseases Ninth and Tenth Revisions (ICD-9/-10) diagnostic codes, pharmacy dispensing records, laboratory results, procedures, vital signs, smoking status, and routinely collected information on alcohol consumption. The VA study was approved by the institutional review boards of Yale University (ref #1506016006) and VA Connecticut Healthcare System (ref #AJ0013) with a waiver of informed consent.

### Exposure groups

Our study comprised three groups: gabapentin initiators, pregabalin initiators, and matched comparators unexposed to either medication. Medication receipt was based on VA outpatient pharmacy dispensing data. We excluded individuals with no outpatient care in the year prior to index date due to the inability to capture baseline data, individuals with no measurement of alcohol consumption in the 2 years prior to index date, and those who reported no alcohol consumption based on the closest measurement to index date. For the gabapentin- and pregabalin-exposed groups, we included all patients who received the medications for at least 60 continuous days for any indication between 1 January 2009 and 30 June 2022, requiring a 180-day washout period from either medication to ensure new exposure episodes. For constructing the unexposed comparator group, we first identified outpatient clinics that were the largest sources of gabapentin and pregabalin prescriptions. We then selected all individuals who attended at least one of these clinics, but did not receive a gabapentin or pregabalin prescription to ensure that unexposed individuals came from the same source population. We randomly selected one visit per unexposed individual to be included in the analyses. Index date was defined as the first dispensed date for gabapentin and pregabalin recipients and the randomly selected outpatient visit date for unexposed individuals.

### Covariates

Data on multiple potential confounders were extracted, including age, race, ethnicity, sex, urban/rural residence, Area Deprivation Index ^45^, geographic region, year of index date, history of clinical conditions and procedures in the two years prior to index date (i.e., AUD, asthma, bariatric surgery, cancer, cerebrovascular disease, chronic obstructive pulmonary disease, congestive heart failure, dementia, diabetes, epilepsy, hemiplegia or paraplegia, human immunodeficiency virus (HIV) infection, liver disease, internalizing disorders (mood disorders, anxiety disorders, and stress-related disorders, including post-traumatic stress disorder), multiple sclerosis, myocardial infarction, opioid use disorder, peptic ulcer disease, peripheral vascular disease, renal disease, rheumatic disease, and traumatic brain injury), Charlson Comorbidity Index, and VACS Index. The Charlson Comorbidity Index is a measure of overall comorbidity based on 17 clinical domains ^46,47^. The VACS Index is a summary score that captures physiologic frailty computed using a validated algorithm that incorporates routine laboratory measures ^48,49^. We also extracted data on medication use at index date (i.e., AUD pharmacotherapies, neurocognitively-active medications, and anticholinergic medications), total medication count, and site-level gabapentin and pregabalin prescribing frequency. Neurocognitively-active medications included prescription opioids, antipsychotics, lithium, anticonvulsants (excluding gabapentin and pregabalin), anti-Parkinson’s, antidepressants, sedatives/hypnotics, muscle relaxants, amphetamine derivatives, antihistamines, and antivertigo agents. We also derived variables capturing substance use at index date (i.e., level of alcohol consumption, smoking status), any substance use treatment program visit, vital signs (body mass index, systolic and diastolic blood pressure, numeric rating scale pain score), laboratory findings (i.e., albumin, total cholesterol, HDL cholesterol, glycated hemoglobin, hemoglobin, total bilirubin, triglycerides, white blood cell count, fibrosis-4 (FIB-4) score, and estimated glomerular filtration rate [eGFR]). We created variables denoting whether the index prescription/visit was in primary care, total number of visits to a prescribing clinic, total number of visits to any clinic, and any hospitalization in the two years prior to index date.

### Outcome Measure and Follow-up

Primary outcome was change in alcohol consumption captured by the AUDIT-C. The VA has performed annual AUDIT-C screening on all patients in primary care since 2008 ^50^. The AUDIT-C is a 3-item questionnaire that assesses frequency and quantity of alcohol use and heavy episodic drinking in the past year ^51^. Scores range from 0 to 12, with the likelihood of alcohol-related morbidity and mortality increasing as scores increase ^52–54^. An AUDIT-C score of 0 indicates no current alcohol use, 1-3 suggests lower-risk drinking, 4-7 suggests at-risk drinking, and ≥8 suggests hazardous or binge alcohol use.

Patients were followed from index date until the earliest of the following: 2 years post-index, their last VA visit, death, or 30 June 2024. Additionally, gabapentin and pregabalin recipients were censored at medication discontinuation. To ensure equal follow-up time within matched pairs, unexposed comparators were censored at the total follow-up time of their matched exposed individual. Although evidence of alcohol consumption at baseline (i.e., AUDIT-C score > 0) was an inclusion criterion, availability of a follow-up AUDIT-C was not required for matching eligibility as such a restriction would not translate to an analogous RCT.

### Statistical Analyses

We conducted propensity score matching to balance the distribution of potential confounders across exposure groups. Propensity scores were estimated using separate multivariable logistic regression models for each exposure contrast: gabapentin versus unexposed (contrast A), pregabalin versus unexposed (contrast B), and pregabalin versus gabapentin (contrast C). A missing category was included for 18 of the 60 covariates with missing data used for matching. Under the additional assumption that associations between fully observed covariates and exposure do not differ across missingness patterns, this approach produces unbiased estimates ^55,56^. For each contrast, individuals from each group were matched 1:1 on the logit of the propensity score using a caliper width 0.20 times the standard deviation of the logit of the propensity score in the region of common support and applying a greedy matching algorithm ^57^. Exact matching was performed on AUD diagnosis and baseline AUDIT-C categories. Given the minimization of potential confounding by indication comparing pregabalin initiators to gabapentin initiators (contrast C), we believed *a priori* that this comparison would provide the most robust evidence whether either medication reduced alcohol use.

We calculated absolute standardized mean difference (SMD) to examine balance between each exposure contrast in the unmatched, matched, and final analytic cohorts, with SMDs ≤0.1 indicating balance ^57^, and average pre- and post-index AUDIT-C scores of individuals in the final analytic cohort. Pre-index AUDIT-C scores were those on or closest to but before the index date, within a maximum of 2 years prior. Post-index AUDIT-C scores were those during and closest to the end of follow-up. Multivariable difference-in-difference DiD linear regression models ^58,59^ were used to estimate the differential change in AUDIT-C scores for each exposure contrast. We performed subgroup analyses by AUD diagnosis, baseline AUDIT-C category, and average daily dose category. Average daily doses were categorized based on previous trial and observational evidence as low (<1,200mg for gabapentin, <300mg for pregabalin), moderate (1,200-1,799mg for gabapentin, 300-599mg for pregabalin), and high (≥1,800mg for gabapentin, ≥600mg for pregabalin). Microsoft SQL Server Management Studio v20.2 and SAS Enterprise Guide v8.3 (SAS Institute) were used for data management and analysis, respectively.

## Results

### Sample

The study flow diagram is presented in Figure 1. We identified 592,957 gabapentin initiators, 14,923 pregabalin initiators, and 2,959,006 eligible unexposed comparators who reported any alcohol consumption in the 2 years prior to the index date. After propensity score matching, contrast A (gabapentin vs. unexposed) included 494,611 matched patients per group, contrast B (pregabalin vs. unexposed) comprised 14,915 matched patients per group, and contrast C (pregabalin vs. gabapentin) included 14,923 patients per group. After excluding individuals with no eligible follow-up AUDIT-C, the final analytic cohorts included 177,189 gabapentin initiators and 140,025 unexposed comparators for contrast A (Supplemental Table 1), 5,270 pregabalin initiators and 4,009 unexposed for contrast B (Supplemental Table 2), and 3,611 pregabalin initiators and 3,384 gabapentin initiators for contrast C (Table 1).

**Figure 1.**
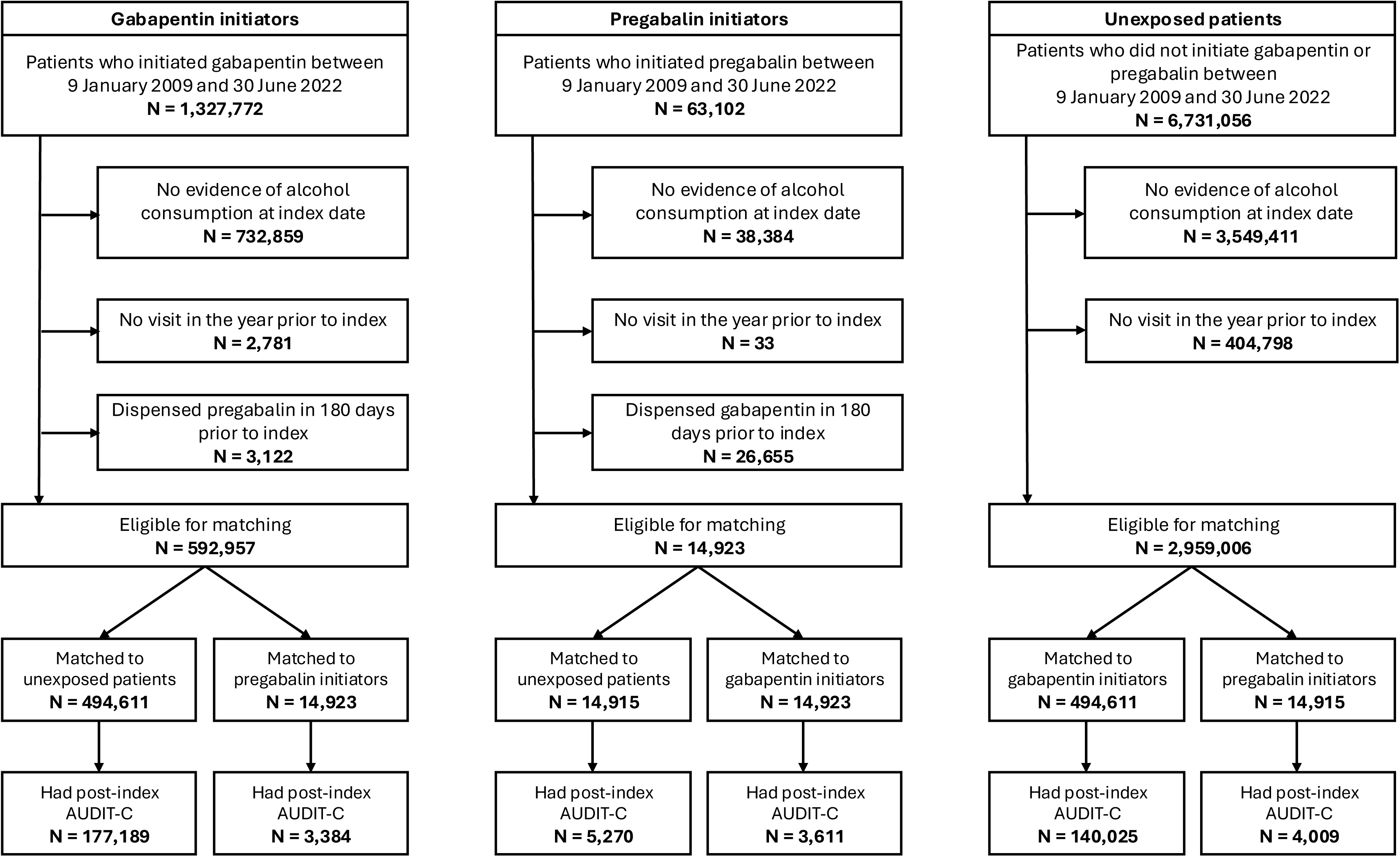
Study flow diagram.

**Table 1.**
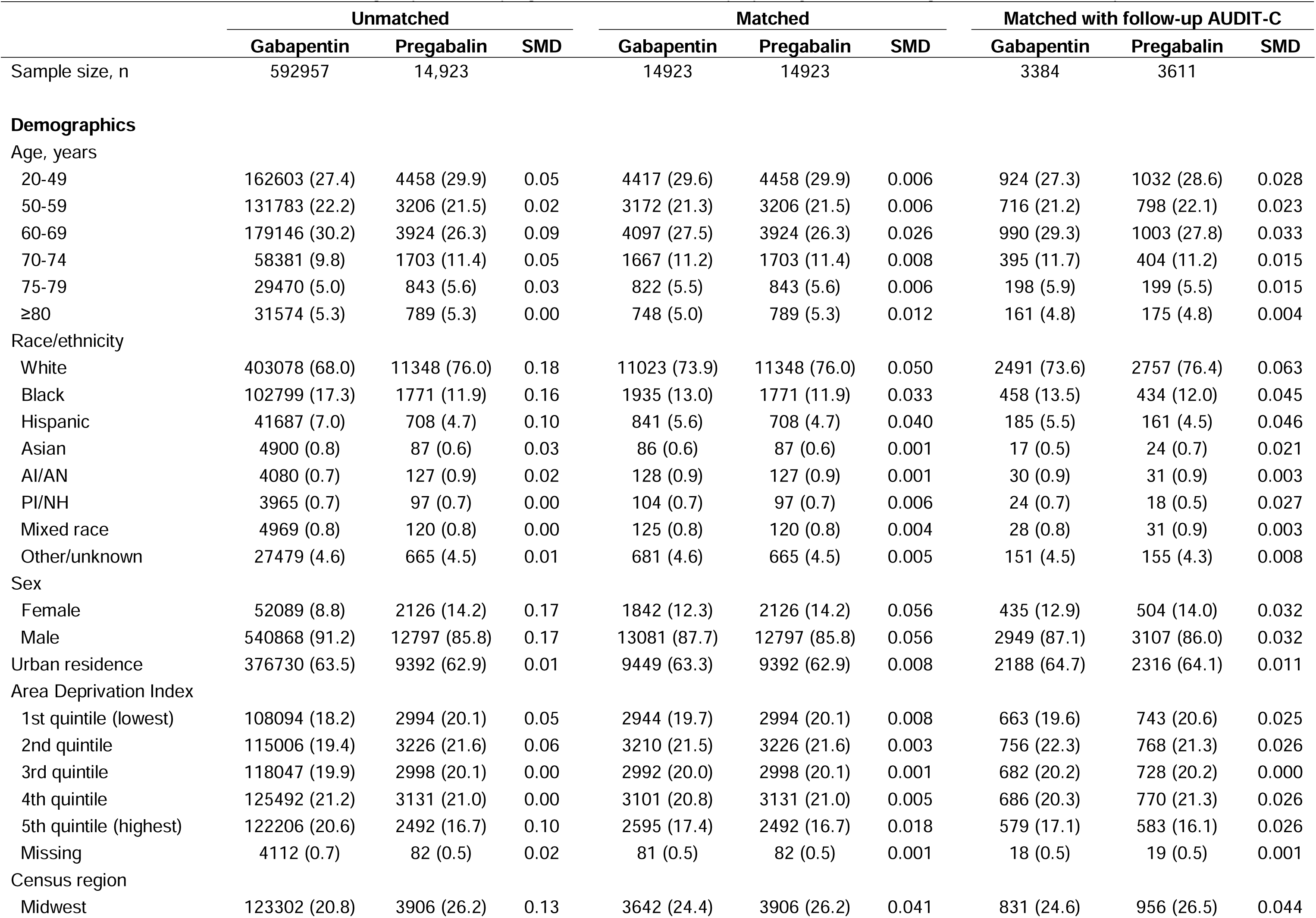
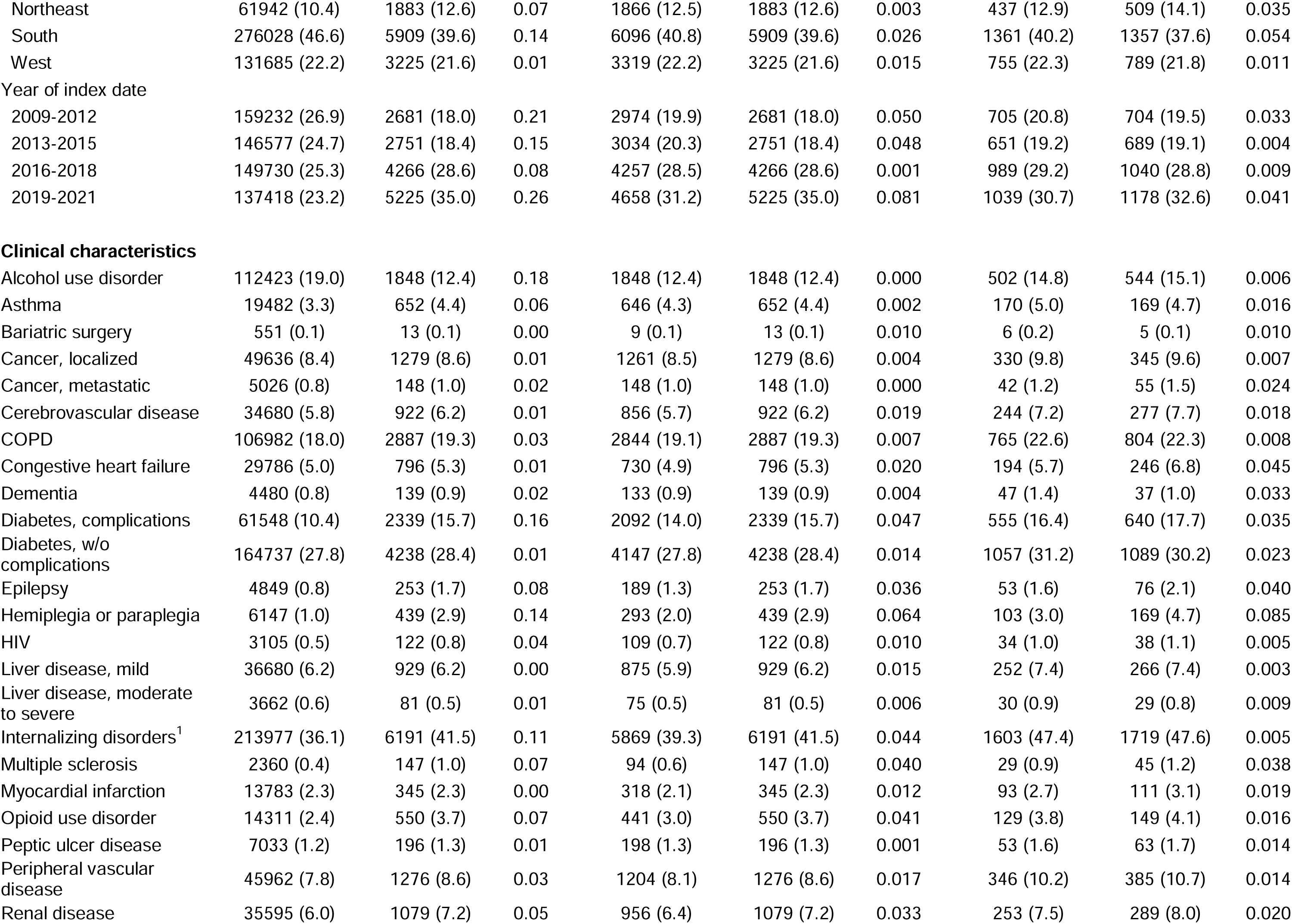
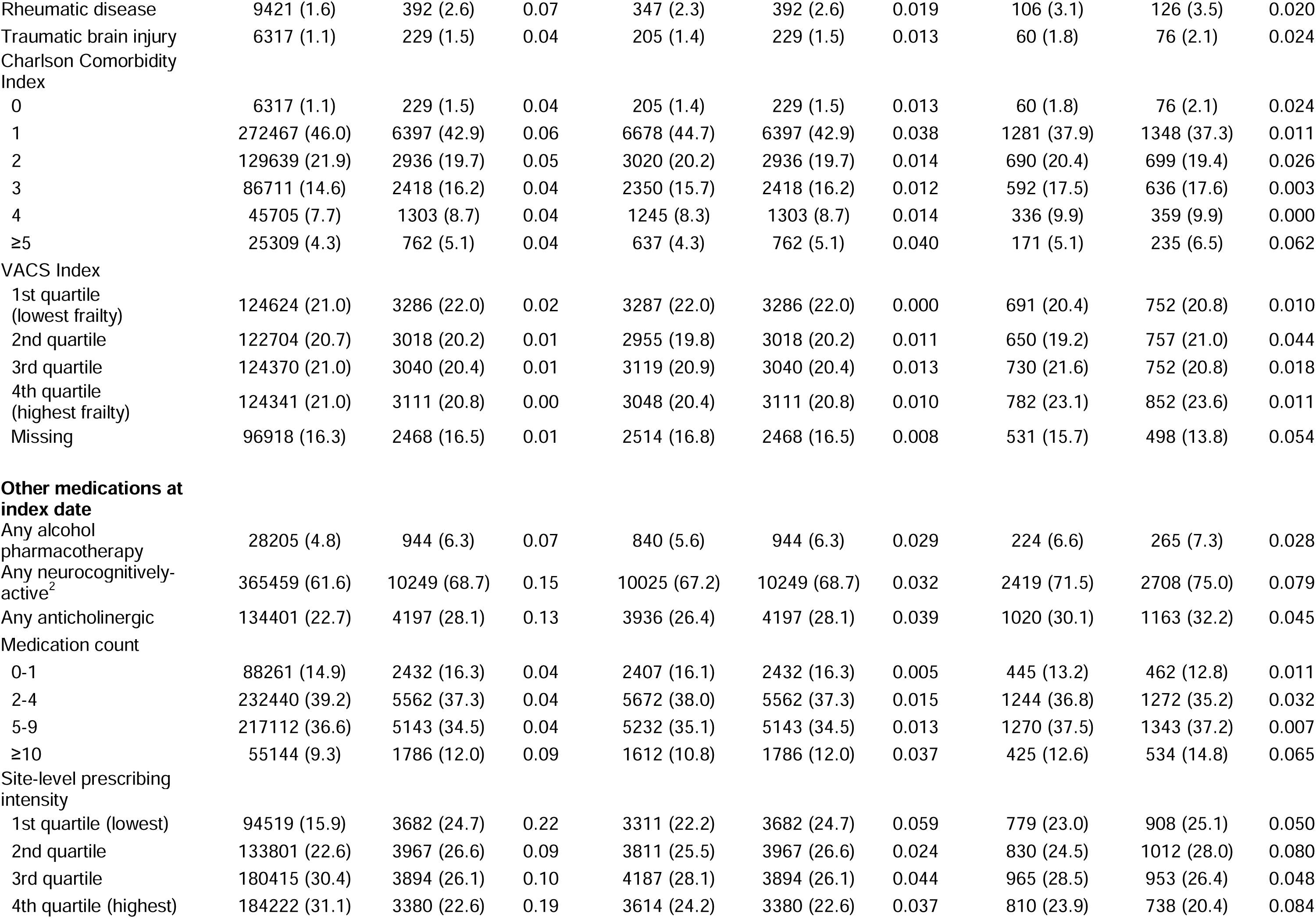
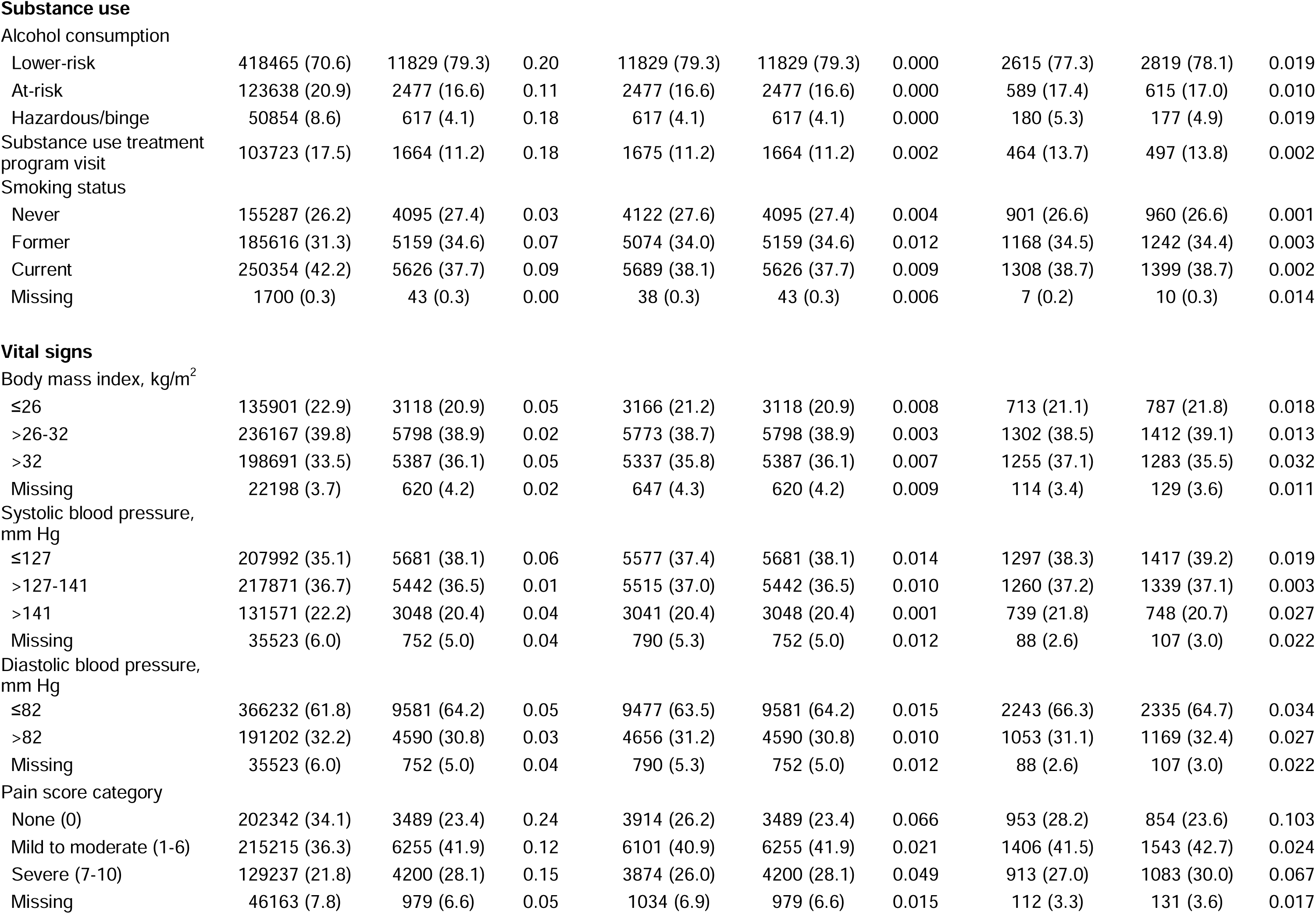
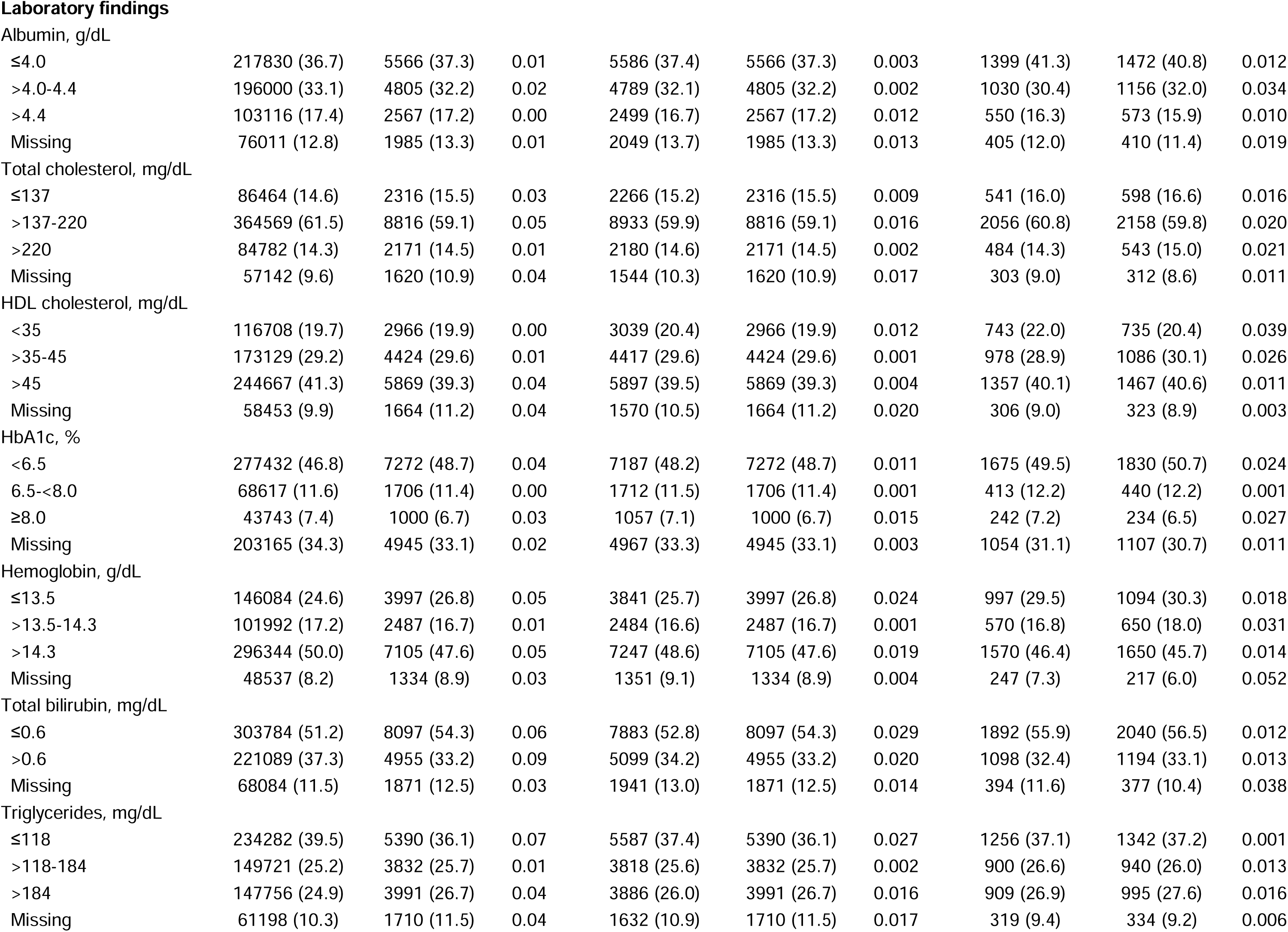
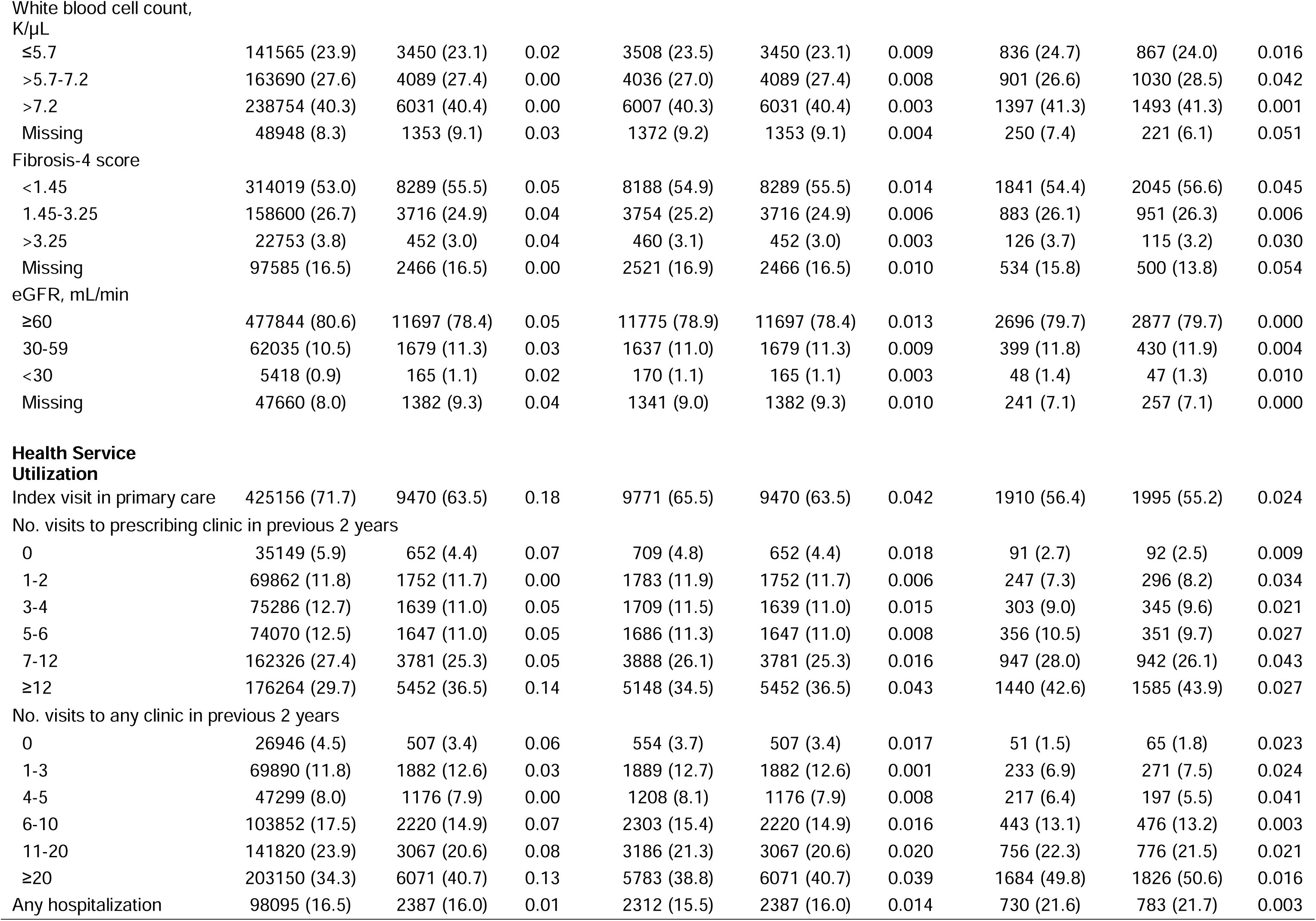
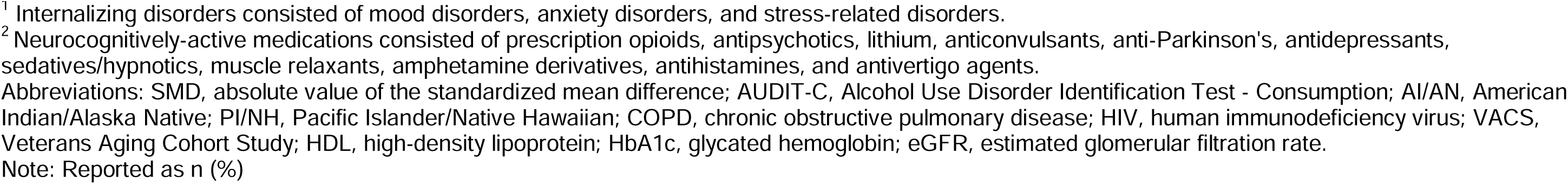
Characteristics between initiators of gabapentin and pregabalin, before and after propensity score matching and recorded follow-up AUDIT-C.

|  | Unmatched |  |  | Matched |  |  | Matched with follow-up AUDIT-C |  |  |
| --- | --- | --- | --- | --- | --- | --- | --- | --- | --- |
|  | Gabapentin | Pregabalin | SMD | Gabapentin | Pregabalin | SMD | Gabapentin | Pregabalin | SMD |
| Sample size, n | 592957 | 14,923 |  | 14923 | 14923 |  | 3384 | 3611 |  |
| **Demographics** |  |  |  |  |  |  |  |  |  |
| Age, years |  |  |  |  |  |  |  |  |  |
| 20-49 | 162603 (27.4) | 4458 (29.9) | 0.05 | 4417 (29.6) | 4458 (29.9) | 0.006 | 924 (27.3) | 1032 (28.6) | 0.028 |
| 50-59 | 131783 (22.2) | 3206 (21.5) | 0.02 | 3172 (21.3) | 3206 (21.5) | 0.006 | 716 (21.2) | 798 (22.1) | 0.023 |
| 60-69 | 179146 (30.2) | 3924 (26.3) | 0.09 | 4097 (27.5) | 3924 (26.3) | 0.026 | 990 (29.3) | 1003 (27.8) | 0.033 |
| 70-74 | 58381 (9.8) | 1703 (11.4) | 0.05 | 1667 (11.2) | 1703 (11.4) | 0.008 | 395 (11.7) | 404 (11.2) | 0.015 |
| 75-79 | 29470 (5.0) | 843 (5.6) | 0.03 | 822 (5.5) | 843 (5.6) | 0.006 | 198 (5.9) | 199 (5.5) | 0.015 |
| ≥80 | 31574 (5.3) | 789 (5.3) | 0.00 | 748 (5.0) | 789 (5.3) | 0.012 | 161 (4.8) | 175 (4.8) | 0.004 |
| Race/ethnicity |  |  |  |  |  |  |  |  |  |
| White | 403078 (68.0) | 11348 (76.0) | 0.18 | 11023 (73.9) | 11348 (76.0) | 0.050 | 2491 (73.6) | 2757 (76.4) | 0.063 |
| Black | 102799 (17.3) | 1771 (11.9) | 0.16 | 1935 (13.0) | 1771 (11.9) | 0.033 | 458 (13.5) | 434 (12.0) | 0.045 |
| Hispanic | 41687 (7.0) | 708 (4.7) | 0.10 | 841 (5.6) | 708 (4.7) | 0.040 | 185 (5.5) | 161 (4.5) | 0.046 |
| Asian | 4900 (0.8) | 87 (0.6) | 0.03 | 86 (0.6) | 87 (0.6) | 0.001 | 17 (0.5) | 24 (0.7) | 0.021 |
| AI/AN | 4080 (0.7) | 127 (0.9) | 0.02 | 128 (0.9) | 127 (0.9) | 0.001 | 30 (0.9) | 31 (0.9) | 0.003 |
| PI/NH | 3965 (0.7) | 97 (0.7) | 0.00 | 104 (0.7) | 97 (0.7) | 0.006 | 24 (0.7) | 18 (0.5) | 0.027 |
| Mixed race | 4969 (0.8) | 120 (0.8) | 0.00 | 125 (0.8) | 120 (0.8) | 0.004 | 28 (0.8) | 31 (0.9) | 0.003 |
| Other/unknown | 27479 (4.6) | 665 (4.5) | 0.01 | 681 (4.6) | 665 (4.5) | 0.005 | 151 (4.5) | 155 (4.3) | 0.008 |
| Sex |  |  |  |  |  |  |  |  |  |
| Female | 52089 (8.8) | 2126 (14.2) | 0.17 | 1842 (12.3) | 2126 (14.2) | 0.056 | 435 (12.9) | 504 (14.0) | 0.032 |
| Male | 540868 (91.2) | 12797 (85.8) | 0.17 | 13081 (87.7) | 12797 (85.8) | 0.056 | 2949 (87.1) | 3107 (86.0) | 0.032 |
| Urban residence | 376730 (63.5) | 9392 (62.9) | 0.01 | 9449 (63.3) | 9392 (62.9) | 0.008 | 2188 (64.7) | 2316 (64.1) | 0.011 |
| Area Deprivation Index |  |  |  |  |  |  |  |  |  |
| 1st quintile (lowest) | 108094 (18.2) | 2994 (20.1) | 0.05 | 2944 (19.7) | 2994 (20.1) | 0.008 | 663 (19.6) | 743 (20.6) | 0.025 |
| 2nd quintile | 115006 (19.4) | 3226 (21.6) | 0.06 | 3210 (21.5) | 3226 (21.6) | 0.003 | 756 (22.3) | 768 (21.3) | 0.026 |
| 3rd quintile | 118047 (19.9) | 2998 (20.1) | 0.00 | 2992 (20.0) | 2998 (20.1) | 0.001 | 682 (20.2) | 728 (20.2) | 0.000 |
| 4th quintile | 125492 (21.2) | 3131 (21.0) | 0.00 | 3101 (20.8) | 3131 (21.0) | 0.005 | 686 (20.3) | 770 (21.3) | 0.026 |
| 5th quintile (highest) | 122206 (20.6) | 2492 (16.7) | 0.10 | 2595 (17.4) | 2492 (16.7) | 0.018 | 579 (17.1) | 583 (16.1) | 0.026 |
| Missing | 4112 (0.7) | 82 (0.5) | 0.02 | 81 (0.5) | 82 (0.5) | 0.001 | 18 (0.5) | 19 (0.5) | 0.001 |
| Census region |  |  |  |  |  |  |  |  |  |
| Midwest | 123302 (20.8) | 3906 (26.2) | 0.13 | 3642 (24.4) | 3906 (26.2) | 0.041 | 831 (24.6) | 956 (26.5) | 0.044 |
| Northeast | 61942 (10.4) | 1883 (12.6) | 0.07 | 1866 (12.5) | 1883 (12.6) | 0.003 | 437 (12.9) | 509 (14.1) | 0.035 |
| South | 276028 (46.6) | 5909 (39.6) | 0.14 | 6096 (40.8) | 5909 (39.6) | 0.026 | 1361 (40.2) | 1357 (37.6) | 0.054 |
| West | 131685 (22.2) | 3225 (21.6) | 0.01 | 3319 (22.2) | 3225 (21.6) | 0.015 | 755 (22.3) | 789 (21.8) | 0.011 |
| Year of index date |  |  |  |  |  |  |  |  |  |
| 2009-2012 | 159232 (26.9) | 2681 (18.0) | 0.21 | 2974 (19.9) | 2681 (18.0) | 0.050 | 705 (20.8) | 704 (19.5) | 0.033 |
| 2013-2015 | 146577 (24.7) | 2751 (18.4) | 0.15 | 3034 (20.3) | 2751 (18.4) | 0.048 | 651 (19.2) | 689 (19.1) | 0.004 |
| 2016-2018 | 149730 (25.3) | 4266 (28.6) | 0.08 | 4257 (28.5) | 4266 (28.6) | 0.001 | 989 (29.2) | 1040 (28.8) | 0.009 |
| 2019-2021 | 137418 (23.2) | 5225 (35.0) | 0.26 | 4658 (31.2) | 5225 (35.0) | 0.081 | 1039 (30.7) | 1178 (32.6) | 0.041 |
| Clinical characteristics |  |  |  |  |  |  |  |  |  |
| Alcohol use disorder | 112423 (19.0) | 1848 (12.4) | 0.18 | 1848 (12.4) | 1848 (12.4) | 0.000 | 502 (14.8) | 544 (15.1) | 0.006 |
| Asthma | 19482 (3.3) | 652 (4.4) | 0.06 | 646 (4.3) | 652 (4.4) | 0.002 | 170 (5.0) | 169 (4.7) | 0.016 |
| Bariatric surgery | 551 (0.1) | 13 (0.1) | 0.00 | 9 (0.1) | 13 (0.1) | 0.010 | 6 (0.2) | 5 (0.1) | 0.010 |
| Cancer, localized | 49636 (8.4) | 1279 (8.6) | 0.01 | 1261 (8.5) | 1279 (8.6) | 0.004 | 330 (9.8) | 345 (9.6) | 0.007 |
| Cancer, metastatic | 5026 (0.8) | 148 (1.0) | 0.02 | 148 (1.0) | 148 (1.0) | 0.000 | 42 (1.2) | 55 (1.5) | 0.024 |
| Cerebrovascular disease | 34680 (5.8) | 922 (6.2) | 0.01 | 856 (5.7) | 922 (6.2) | 0.019 | 244 (7.2) | 277 (7.7) | 0.018 |
| COPD | 106982 (18.0) | 2887 (19.3) | 0.03 | 2844 (19.1) | 2887 (19.3) | 0.007 | 765 (22.6) | 804 (22.3) | 0.008 |
| Congestive heart failure | 29786 (5.0) | 796 (5.3) | 0.01 | 730 (4.9) | 796 (5.3) | 0.020 | 194 (5.7) | 246 (6.8) | 0.045 |
| Dementia | 4480 (0.8) | 139 (0.9) | 0.02 | 133 (0.9) | 139 (0.9) | 0.004 | 47 (1.4) | 37 (1.0) | 0.033 |
| Diabetes, complications | 61548 (10.4) | 2339 (15.7) | 0.16 | 2092 (14.0) | 2339 (15.7) | 0.047 | 555 (16.4) | 640 (17.7) | 0.035 |
| Diabetes, w/o complications | 164737 (27.8) | 4238 (28.4) | 0.01 | 4147 (27.8) | 4238 (28.4) | 0.014 | 1057 (31.2) | 1089 (30.2) | 0.023 |
| Epilepsy | 4849 (0.8) | 253 (1.7) | 0.08 | 189 (1.3) | 253 (1.7) | 0.036 | 53 (1.6) | 76 (2.1) | 0.040 |
| Hemiplegia or paraplegia | 6147 (1.0) | 439 (2.9) | 0.14 | 293 (2.0) | 439 (2.9) | 0.064 | 103 (3.0) | 169 (4.7) | 0.085 |
| HIV | 3105 (0.5) | 122 (0.8) | 0.04 | 109 (0.7) | 122 (0.8) | 0.010 | 34 (1.0) | 38 (1.1) | 0.005 |
| Liver disease, mild | 36680 (6.2) | 929 (6.2) | 0.00 | 875 (5.9) | 929 (6.2) | 0.015 | 252 (7.4) | 266 (7.4) | 0.003 |
| Liver disease, moderate to severe | 3662 (0.6) | 81 (0.5) | 0.01 | 75 (0.5) | 81 (0.5) | 0.006 | 30 (0.9) | 29 (0.8) | 0.009 |
| Internalizing disorders<sup>1</sup> | 213977 (36.1) | 6191 (41.5) | 0.11 | 5869 (39.3) | 6191 (41.5) | 0.044 | 1603 (47.4) | 1719 (47.6) | 0.005 |
| Multiple sclerosis | 2360 (0.4) | 147 (1.0) | 0.07 | 94 (0.6) | 147 (1.0) | 0.040 | 29 (0.9) | 45 (1.2) | 0.038 |
| Myocardial infarction | 13783 (2.3) | 345 (2.3) | 0.00 | 318 (2.1) | 345 (2.3) | 0.012 | 93 (2.7) | 111 (3.1) | 0.019 |
| Opioid use disorder | 14311 (2.4) | 550 (3.7) | 0.07 | 441 (3.0) | 550 (3.7) | 0.041 | 129 (3.8) | 149 (4.1) | 0.016 |
| Peptic ulcer disease | 7033 (1.2) | 196 (1.3) | 0.01 | 198 (1.3) | 196 (1.3) | 0.001 | 53 (1.6) | 63 (1.7) | 0.014 |
| Peripheral vascular disease | 45962 (7.8) | 1276 (8.6) | 0.03 | 1204 (8.1) | 1276 (8.6) | 0.017 | 346 (10.2) | 385 (10.7) | 0.014 |
| Renal disease | 35595 (6.0) | 1079 (7.2) | 0.05 | 956 (6.4) | 1079 (7.2) | 0.033 | 253 (7.5) | 289 (8.0) | 0.020 |
| Rheumatic disease | 9421 (1.6) | 392 (2.6) | 0.07 | 347 (2.3) | 392 (2.6) | 0.019 | 106 (3.1) | 126 (3.5) | 0.020 |
| Traumatic brain injury | 6317 (1.1) | 229 (1.5) | 0.04 | 205 (1.4) | 229 (1.5) | 0.013 | 60 (1.8) | 76 (2.1) | 0.024 |
| Charlson Comorbidity Index |  |  |  |  |  |  |  |  |  |
| 0 | 6317 (1.1) | 229 (1.5) | 0.04 | 205 (1.4) | 229 (1.5) | 0.013 | 60 (1.8) | 76 (2.1) | 0.024 |
| 1 | 272467 (46.0) | 6397 (42.9) | 0.06 | 6678 (44.7) | 6397 (42.9) | 0.038 | 1281 (37.9) | 1348 (37.3) | 0.011 |
| 2 | 129639 (21.9) | 2936 (19.7) | 0.05 | 3020 (20.2) | 2936 (19.7) | 0.014 | 690 (20.4) | 699 (19.4) | 0.026 |
| 3 | 86711 (14.6) | 2418 (16.2) | 0.04 | 2350 (15.7) | 2418 (16.2) | 0.012 | 592 (17.5) | 636 (17.6) | 0.003 |
| 4 | 45705 (7.7) | 1303 (8.7) | 0.04 | 1245 (8.3) | 1303 (8.7) | 0.014 | 336 (9.9) | 359 (9.9) | 0.000 |
| ≥5 | 25309 (4.3) | 762 (5.1) | 0.04 | 637 (4.3) | 762 (5.1) | 0.040 | 171 (5.1) | 235 (6.5) | 0.062 |
| VACS Index |  |  |  |  |  |  |  |  |  |
| 1st quartile (lowest frailty) | 124624 (21.0) | 3286 (22.0) | 0.02 | 3287 (22.0) | 3286 (22.0) | 0.000 | 691 (20.4) | 752 (20.8) | 0.010 |
| 2nd quartile | 122704 (20.7) | 3018 (20.2) | 0.01 | 2955 (19.8) | 3018 (20.2) | 0.011 | 650 (19.2) | 757 (21.0) | 0.044 |
| 3rd quartile | 124370 (21.0) | 3040 (20.4) | 0.01 | 3119 (20.9) | 3040 (20.4) | 0.013 | 730 (21.6) | 752 (20.8) | 0.018 |
| 4th quartile (highest frailty) | 124341 (21.0) | 3111 (20.8) | 0.00 | 3048 (20.4) | 3111 (20.8) | 0.010 | 782 (23.1) | 852 (23.6) | 0.011 |
| Missing | 96918 (16.3) | 2468 (16.5) | 0.01 | 2514 (16.8) | 2468 (16.5) | 0.008 | 531 (15.7) | 498 (13.8) | 0.054 |
| **Other medications at index date** |  |  |  |  |  |  |  |  |  |
| Any alcohol pharmacotherapy | 28205 (4.8) | 944 (6.3) | 0.07 | 840 (5.6) | 944 (6.3) | 0.029 | 224 (6.6) | 265 (7.3) | 0.028 |
| Any neurocognitively-active<sup>2</sup> | 365459 (61.6) | 10249 (68.7) | 0.15 | 10025 (67.2) | 10249 (68.7) | 0.032 | 2419 (71.5) | 2708 (75.0) | 0.079 |
| Any anticholinergic | 134401 (22.7) | 4197 (28.1) | 0.13 | 3936 (26.4) | 4197 (28.1) | 0.039 | 1020 (30.1) | 1163 (32.2) | 0.045 |
| Medication count |  |  |  |  |  |  |  |  |  |
| 0-1 | 88261 (14.9) | 2432 (16.3) | 0.04 | 2407 (16.1) | 2432 (16.3) | 0.005 | 445 (13.2) | 462 (12.8) | 0.011 |
| 2-4 | 232440 (39.2) | 5562 (37.3) | 0.04 | 5672 (38.0) | 5562 (37.3) | 0.015 | 1244 (36.8) | 1272 (35.2) | 0.032 |
| 5-9 | 217112 (36.6) | 5143 (34.5) | 0.04 | 5232 (35.1) | 5143 (34.5) | 0.013 | 1270 (37.5) | 1343 (37.2) | 0.007 |
| ≥10 | 55144 (9.3) | 1786 (12.0) | 0.09 | 1612 (10.8) | 1786 (12.0) | 0.037 | 425 (12.6) | 534 (14.8) | 0.065 |
| Site-level prescribing intensity |  |  |  |  |  |  |  |  |  |
| 1st quartile (lowest) | 94519 (15.9) | 3682 (24.7) | 0.22 | 3311 (22.2) | 3682 (24.7) | 0.059 | 779 (23.0) | 908 (25.1) | 0.050 |
| 2nd quartile | 133801 (22.6) | 3967 (26.6) | 0.09 | 3811 (25.5) | 3967 (26.6) | 0.024 | 830 (24.5) | 1012 (28.0) | 0.080 |
| 3rd quartile | 180415 (30.4) | 3894 (26.1) | 0.10 | 4187 (28.1) | 3894 (26.1) | 0.044 | 965 (28.5) | 953 (26.4) | 0.048 |
| 4th quartile (highest) | 184222 (31.1) | 3380 (22.6) | 0.19 | 3614 (24.2) | 3380 (22.6) | 0.037 | 810 (23.9) | 738 (20.4) | 0.084 |
| Alcohol consumption |  |  |  |  |  |  |  |  |  |
| Lower-risk | 418465 (70.6) | 11829 (79.3) | 0.20 | 11829 (79.3) | 11829 (79.3) | 0.000 | 2615 (77.3) | 2819 (78.1) | 0.019 |
| At-risk | 123638 (20.9) | 2477 (16.6) | 0.11 | 2477 (16.6) | 2477 (16.6) | 0.000 | 589 (17.4) | 615 (17.0) | 0.010 |
| Hazardous/binge | 50854 (8.6) | 617 (4.1) | 0.18 | 617 (4.1) | 617 (4.1) | 0.000 | 180 (5.3) | 177 (4.9) | 0.019 |
| Substance use treatment program visit | 103723 (17.5) | 1664 (11.2) | 0.18 | 1675 (11.2) | 1664 (11.2) | 0.002 | 464 (13.7) | 497 (13.8) | 0.002 |
| Smoking status |  |  |  |  |  |  |  |  |  |
| Never | 155287 (26.2) | 4095 (27.4) | 0.03 | 4122 (27.6) | 4095 (27.4) | 0.004 | 901 (26.6) | 960 (26.6) | 0.001 |
| Former | 185616 (31.3) | 5159 (34.6) | 0.07 | 5074 (34.0) | 5159 (34.6) | 0.012 | 1168 (34.5) | 1242 (34.4) | 0.003 |
| Current | 250354 (42.2) | 5626 (37.7) | 0.09 | 5689 (38.1) | 5626 (37.7) | 0.009 | 1308 (38.7) | 1399 (38.7) | 0.002 |
| Missing | 1700 (0.3) | 43 (0.3) | 0.00 | 38 (0.3) | 43 (0.3) | 0.006 | 7 (0.2) | 10 (0.3) | 0.014 |
| Body mass index, kg/m<sup>2</sup> |  |  |  |  |  |  |  |  |  |
| ≤26 | 135901 (22.9) | 3118 (20.9) | 0.05 | 3166 (21.2) | 3118 (20.9) | 0.008 | 713 (21.1) | 787 (21.8) | 0.018 |
| >26-32 | 236167 (39.8) | 5798 (38.9) | 0.02 | 5773 (38.7) | 5798 (38.9) | 0.003 | 1302 (38.5) | 1412 (39.1) | 0.013 |
| >32 | 198691 (33.5) | 5387 (36.1) | 0.05 | 5337 (35.8) | 5387 (36.1) | 0.007 | 1255 (37.1) | 1283 (35.5) | 0.032 |
| Missing | 22198 (3.7) | 620 (4.2) | 0.02 | 647 (4.3) | 620 (4.2) | 0.009 | 114 (3.4) | 129 (3.6) | 0.011 |
| Systolic blood pressure, mm Hg |  |  |  |  |  |  |  |  |  |
| ≤127 | 207992 (35.1) | 5681 (38.1) | 0.06 | 5577 (37.4) | 5681 (38.1) | 0.014 | 1297 (38.3) | 1417 (39.2) | 0.019 |
| >127-141 | 217871 (36.7) | 5442 (36.5) | 0.01 | 5515 (37.0) | 5442 (36.5) | 0.010 | 1260 (37.2) | 1339 (37.1) | 0.003 |
| >141 | 131571 (22.2) | 3048 (20.4) | 0.04 | 3041 (20.4) | 3048 (20.4) | 0.001 | 739 (21.8) | 748 (20.7) | 0.027 |
| Missing | 35523 (6.0) | 752 (5.0) | 0.04 | 790 (5.3) | 752 (5.0) | 0.012 | 88 (2.6) | 107 (3.0) | 0.022 |
| Diastolic blood pressure, mm Hg |  |  |  |  |  |  |  |  |  |
| ≤82 | 366232 (61.8) | 9581 (64.2) | 0.05 | 9477 (63.5) | 9581 (64.2) | 0.015 | 2243 (66.3) | 2335 (64.7) | 0.034 |
| >82 | 191202 (32.2) | 4590 (30.8) | 0.03 | 4656 (31.2) | 4590 (30.8) | 0.010 | 1053 (31.1) | 1169 (32.4) | 0.027 |
| Missing | 35523 (6.0) | 752 (5.0) | 0.04 | 790 (5.3) | 752 (5.0) | 0.012 | 88 (2.6) | 107 (3.0) | 0.022 |
| Pain score category |  |  |  |  |  |  |  |  |  |
| None (0) | 202342 (34.1) | 3489 (23.4) | 0.24 | 3914 (26.2) | 3489 (23.4) | 0.066 | 953 (28.2) | 854 (23.6) | 0.103 |
| Mild to moderate (1-6) | 215215 (36.3) | 6255 (41.9) | 0.12 | 6101 (40.9) | 6255 (41.9) | 0.021 | 1406 (41.5) | 1543 (42.7) | 0.024 |
| Severe (7-10) | 129237 (21.8) | 4200 (28.1) | 0.15 | 3874 (26.0) | 4200 (28.1) | 0.049 | 913 (27.0) | 1083 (30.0) | 0.067 |
| Missing | 46163 (7.8) | 979 (6.6) | 0.05 | 1034 (6.9) | 979 (6.6) | 0.015 | 112 (3.3) | 131 (3.6) | 0.017 |
| Albumin, g/dL |  |  |  |  |  |  |  |  |  |
| ≤4.0 | 217830 (36.7) | 5566 (37.3) | 0.01 | 5586 (37.4) | 5566 (37.3) | 0.003 | 1399 (41.3) | 1472 (40.8) | 0.012 |
| >4.0-4.4 | 196000 (33.1) | 4805 (32.2) | 0.02 | 4789 (32.1) | 4805 (32.2) | 0.002 | 1030 (30.4) | 1156 (32.0) | 0.034 |
| >4.4 | 103116 (17.4) | 2567 (17.2) | 0.00 | 2499 (16.7) | 2567 (17.2) | 0.012 | 550 (16.3) | 573 (15.9) | 0.010 |
| Missing | 76011 (12.8) | 1985 (13.3) | 0.01 | 2049 (13.7) | 1985 (13.3) | 0.013 | 405 (12.0) | 410 (11.4) | 0.019 |
| Total cholesterol, mg/dL |  |  |  |  |  |  |  |  |  |
| ≤137 | 86464 (14.6) | 2316 (15.5) | 0.03 | 2266 (15.2) | 2316 (15.5) | 0.009 | 541 (16.0) | 598 (16.6) | 0.016 |
| >137-220 | 364569 (61.5) | 8816 (59.1) | 0.05 | 8933 (59.9) | 8816 (59.1) | 0.016 | 2056 (60.8) | 2158 (59.8) | 0.020 |
| >220 | 84782 (14.3) | 2171 (14.5) | 0.01 | 2180 (14.6) | 2171 (14.5) | 0.002 | 484 (14.3) | 543 (15.0) | 0.021 |
| Missing | 57142 (9.6) | 1620 (10.9) | 0.04 | 1544 (10.3) | 1620 (10.9) | 0.017 | 303 (9.0) | 312 (8.6) | 0.011 |
| HDL cholesterol, mg/dL |  |  |  |  |  |  |  |  |  |
| <35 | 116708 (19.7) | 2966 (19.9) | 0.00 | 3039 (20.4) | 2966 (19.9) | 0.012 | 743 (22.0) | 735 (20.4) | 0.039 |
| >35-45 | 173129 (29.2) | 4424 (29.6) | 0.01 | 4417 (29.6) | 4424 (29.6) | 0.001 | 978 (28.9) | 1086 (30.1) | 0.026 |
| >45 | 244667 (41.3) | 5869 (39.3) | 0.04 | 5897 (39.5) | 5869 (39.3) | 0.004 | 1357 (40.1) | 1467 (40.6) | 0.011 |
| Missing | 58453 (9.9) | 1664 (11.2) | 0.04 | 1570 (10.5) | 1664 (11.2) | 0.020 | 306 (9.0) | 323 (8.9) | 0.003 |
| HbA1c, % |  |  |  |  |  |  |  |  |  |
| <6.5 | 277432 (46.8) | 7272 (48.7) | 0.04 | 7187 (48.2) | 7272 (48.7) | 0.011 | 1675 (49.5) | 1830 (50.7) | 0.024 |
| 6.5-<8.0 | 68617 (11.6) | 1706 (11.4) | 0.00 | 1712 (11.5) | 1706 (11.4) | 0.001 | 413 (12.2) | 440 (12.2) | 0.001 |
| ≥8.0 | 43743 (7.4) | 1000 (6.7) | 0.03 | 1057 (7.1) | 1000 (6.7) | 0.015 | 242 (7.2) | 234 (6.5) | 0.027 |
| Missing | 203165 (34.3) | 4945 (33.1) | 0.02 | 4967 (33.3) | 4945 (33.1) | 0.003 | 1054 (31.1) | 1107 (30.7) | 0.011 |
| Hemoglobin, g/dL |  |  |  |  |  |  |  |  |  |
| ≤13.5 | 146084 (24.6) | 3997 (26.8) | 0.05 | 3841 (25.7) | 3997 (26.8) | 0.024 | 997 (29.5) | 1094 (30.3) | 0.018 |
| >13.5-14.3 | 101992 (17.2) | 2487 (16.7) | 0.01 | 2484 (16.6) | 2487 (16.7) | 0.001 | 570 (16.8) | 650 (18.0) | 0.031 |
| >14.3 | 296344 (50.0) | 7105 (47.6) | 0.05 | 7247 (48.6) | 7105 (47.6) | 0.019 | 1570 (46.4) | 1650 (45.7) | 0.014 |
| Missing | 48537 (8.2) | 1334 (8.9) | 0.03 | 1351 (9.1) | 1334 (8.9) | 0.004 | 247 (7.3) | 217 (6.0) | 0.052 |
| Total bilirubin, mg/dL |  |  |  |  |  |  |  |  |  |
| ≤0.6 | 303784 (51.2) | 8097 (54.3) | 0.06 | 7883 (52.8) | 8097 (54.3) | 0.029 | 1892 (55.9) | 2040 (56.5) | 0.012 |
| >0.6 | 221089 (37.3) | 4955 (33.2) | 0.09 | 5099 (34.2) | 4955 (33.2) | 0.020 | 1098 (32.4) | 1194 (33.1) | 0.013 |
| Missing | 68084 (11.5) | 1871 (12.5) | 0.03 | 1941 (13.0) | 1871 (12.5) | 0.014 | 394 (11.6) | 377 (10.4) | 0.038 |
| Triglycerides, mg/dL |  |  |  |  |  |  |  |  |  |
| ≤118 | 234282 (39.5) | 5390 (36.1) | 0.07 | 5587 (37.4) | 5390 (36.1) | 0.027 | 1256 (37.1) | 1342 (37.2) | 0.001 |
| >118-184 | 149721 (25.2) | 3832 (25.7) | 0.01 | 3818 (25.6) | 3832 (25.7) | 0.002 | 900 (26.6) | 940 (26.0) | 0.013 |
| >184 | 147756 (24.9) | 3991 (26.7) | 0.04 | 3886 (26.0) | 3991 (26.7) | 0.016 | 909 (26.9) | 995 (27.6) | 0.016 |
| Missing | 61198 (10.3) | 1710 (11.5) | 0.04 | 1632 (10.9) | 1710 (11.5) | 0.017 | 319 (9.4) | 334 (9.2) | 0.006 |
| White blood cell count, K/μL | | | | | | | | | |
| ≤5.7 | 141565 (23.9) | 3450 (23.1) | 0.02 | 3508 (23.5) | 3450 (23.1) | 0.009 | 836 (24.7) | 867 (24.0) | 0.016 |
| >5.7-7.2 | 163690 (27.6) | 4089 (27.4) | 0.00 | 4036 (27.0) | 4089 (27.4) | 0.008 | 901 (26.6) | 1030 (28.5) | 0.042 |
| >7.2 | 238754 (40.3) | 6031 (40.4) | 0.00 | 6007 (40.3) | 6031 (40.4) | 0.003 | 1397 (41.3) | 1493 (41.3) | 0.001 |
| Missing | 48948 (8.3) | 1353 (9.1) | 0.03 | 1372 (9.2) | 1353 (9.1) | 0.004 | 250 (7.4) | 221 (6.1) | 0.051 |
| Fibrosis-4 score |  |  |  |  |  |  |  |  |  |
| <1.45 | 314019 (53.0) | 8289 (55.5) | 0.05 | 8188 (54.9) | 8289 (55.5) | 0.014 | 1841 (54.4) | 2045 (56.6) | 0.045 |
| 1.45-3.25 | 158600 (26.7) | 3716 (24.9) | 0.04 | 3754 (25.2) | 3716 (24.9) | 0.006 | 883 (26.1) | 951 (26.3) | 0.006 |
| >3.25 | 22753 (3.8) | 452 (3.0) | 0.04 | 460 (3.1) | 452 (3.0) | 0.003 | 126 (3.7) | 115 (3.2) | 0.030 |
| Missing | 97585 (16.5) | 2466 (16.5) | 0.00 | 2521 (16.9) | 2466 (16.5) | 0.010 | 534 (15.8) | 500 (13.8) | 0.054 |
| eGFR, mL/min |  |  |  |  |  |  |  |  |  |
| ≥60 | 477844 (80.6) | 11697 (78.4) | 0.05 | 11775 (78.9) | 11697 (78.4) | 0.013 | 2696 (79.7) | 2877 (79.7) | 0.000 |
| 30-59 | 62035 (10.5) | 1679 (11.3) | 0.03 | 1637 (11.0) | 1679 (11.3) | 0.009 | 399 (11.8) | 430 (11.9) | 0.004 |
| <30 | 5418 (0.9) | 165 (1.1) | 0.02 | 170 (1.1) | 165 (1.1) | 0.003 | 48 (1.4) | 47 (1.3) | 0.010 |
| Missing | 47660 (8.0) | 1382 (9.3) | 0.04 | 1341 (9.0) | 1382 (9.3) | 0.010 | 241 (7.1) | 257 (7.1) | 0.000 |
| **Health Service Utilization** |  |  |  |  |  |  |  |  |  |
| Index visit in primary care | 425156 (71.7) | 9470 (63.5) | 0.18 | 9771 (65.5) | 9470 (63.5) | 0.042 | 1910 (56.4) | 1995 (55.2) | 0.024 |
| No. visits to prescribing clinic in previous 2 years |  |  |  |  |  |  |  |  |  |
| 0 | 35149 (5.9) | 652 (4.4) | 0.07 | 709 (4.8) | 652 (4.4) | 0.018 | 91 (2.7) | 92 (2.5) | 0.009 |
| 1-2 | 69862 (11.8) | 1752 (11.7) | 0.00 | 1783 (11.9) | 1752 (11.7) | 0.006 | 247 (7.3) | 296 (8.2) | 0.034 |
| 3-4 | 75286 (12.7) | 1639 (11.0) | 0.05 | 1709 (11.5) | 1639 (11.0) | 0.015 | 303 (9.0) | 345 (9.6) | 0.021 |
| 5-6 | 74070 (12.5) | 1647 (11.0) | 0.05 | 1686 (11.3) | 1647 (11.0) | 0.008 | 356 (10.5) | 351 (9.7) | 0.027 |
| 7-12 | 162326 (27.4) | 3781 (25.3) | 0.05 | 3888 (26.1) | 3781 (25.3) | 0.016 | 947 (28.0) | 942 (26.1) | 0.043 |
| ≥12 | 176264 (29.7) | 5452 (36.5) | 0.14 | 5148 (34.5) | 5452 (36.5) | 0.043 | 1440 (42.6) | 1585 (43.9) | 0.027 |
| No. visits to any clinic in previous 2 years |  |  |  |  |  |  |  |  |  |
| 0 | 26946 (4.5) | 507 (3.4) | 0.06 | 554 (3.7) | 507 (3.4) | 0.017 | 51 (1.5) | 65 (1.8) | 0.023 |
| 1-3 | 69890 (11.8) | 1882 (12.6) | 0.03 | 1889 (12.7) | 1882 (12.6) | 0.001 | 233 (6.9) | 271 (7.5) | 0.024 |
| 4-5 | 47299 (8.0) | 1176 (7.9) | 0.00 | 1208 (8.1) | 1176 (7.9) | 0.008 | 217 (6.4) | 197 (5.5) | 0.041 |
| 6-10 | 103852 (17.5) | 2220 (14.9) | 0.07 | 2303 (15.4) | 2220 (14.9) | 0.016 | 443 (13.1) | 476 (13.2) | 0.003 |
| 11-20 | 141820 (23.9) | 3067 (20.6) | 0.08 | 3186 (21.3) | 3067 (20.6) | 0.020 | 756 (22.3) | 776 (21.5) | 0.021 |
| ≥20 | 203150 (34.3) | 6071 (40.7) | 0.13 | 5783 (38.8) | 6071 (40.7) | 0.039 | 1684 (49.8) | 1826 (50.6) | 0.016 |
| Any hospitalization | 98095 (16.5) | 2387 (16.0) | 0.01 | 2312 (15.5) | 2387 (16.0) | 0.014 | 730 (21.6) | 783 (21.7) | 0.003 |

<sup>1</sup> Internalizing disorders consisted of mood disorders, anxiety disorders, and stress-related disorders.
<sup>2</sup> Neurocognitively-active medications consisted of prescription opioids, antipsychotics, lithium, anticonvulsants, anti-Parkinson's, antidepressants, sedatives/hypnotics, muscle relaxants, amphetamine derivatives, antihistamines, and antivertigo agents.
Abbreviations: SMD, absolute value of the standardized mean difference; AUDIT-C, Alcohol Use Disorder Identification Test - Consumption; AI/AN, American Indian/Alaska Native; PI/NH, Pacific Islander/Native Hawaiian; COPD, chronic obstructive pulmonary disease; HIV, human immunodeficiency virus; VACS, Veterans Aging Cohort Study; HDL, high-density lipoprotein; HbA1c, glycated hemoglobin; eGFR, estimated glomerular filtration rate.
Note: Reported as n (%)

Distribution of baseline characteristics differed between groups before propensity score matching. After propensity score matching and restricting patients to those with a follow-up AUDIT-C, the distribution of baseline characteristics was balanced between groups (nearly all SMDs≤0.1 except for six variables with SMDs≤0.16 for contrast A [Supplemental Table S1], and three variables with SMD≤0.11 for contrast B [Supplemental Table S2]). The distribution of propensity scores for each contrast before and after matching are shown in Supplemental Figure S1.

### Changes in alcohol consumption – Contrast A (Gabapentin vs. Unexposed)

Pre-index and post-index AUDIT-C scores among gabapentin initiators was 2.93 (standard error [SE]=0.01) and 2.20 (SE=0.01), respectively, resulting in a difference of −0.72 (SE=0.01). Pre-index and post-index AUDIT-C scores among unexposed comparators was 2.83 (SE=0.01) and 2.19 (SE=0.01), respectively, resulting in a difference of −0.63 (SE=0.01). Gabapentin initiators had a significantly greater reduction in AUDIT-C scores than unexposed comparators (DiD: 0.09 points, 95% CI: 0.06, 0.11, p<0.0001; Table 2, Figure 2A). This effect was consistent in subgroup analyses based on diagnosed AUD and by baseline AUDIT-C category (Table 2, Figure 2A). A dose-response relationship was observed, with a small effect at low average daily doses (DiD: 0.04 points, 95% CI: 0.01, 0.07, p=0.0027), an effect consistent to the overall estimate at moderate doses (DiD: 0.11 points, 95% CI: 0.06, 0.16, p<0.0001), and the largest effect at high doses (DiD: 0.22 points, 95% CI: 0.18, 0.26, p<0.0001).

**Figure 2.**
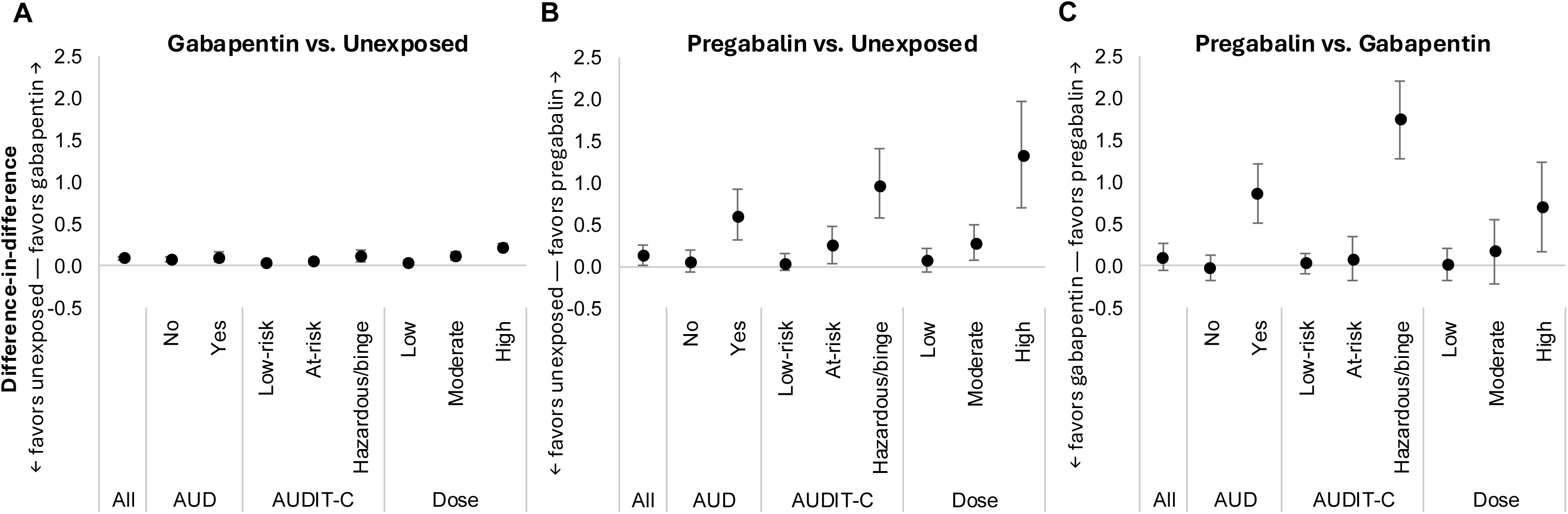
Association between receipt of gabapentin and pregabalin and alcohol use. Difference-in-difference estimates and 95% confidence intervals of changes in AUDIT-C scores stratified by baseline AUD diagnosis and by baseline AUDIT-C risk categories. (A) Gabapentin recipients vs. unexposed individuals, (B) pregabalin recipients vs. unexposed individuals, (C) pregabalin recipients vs. gabapentin recipients.

**Table 2.**
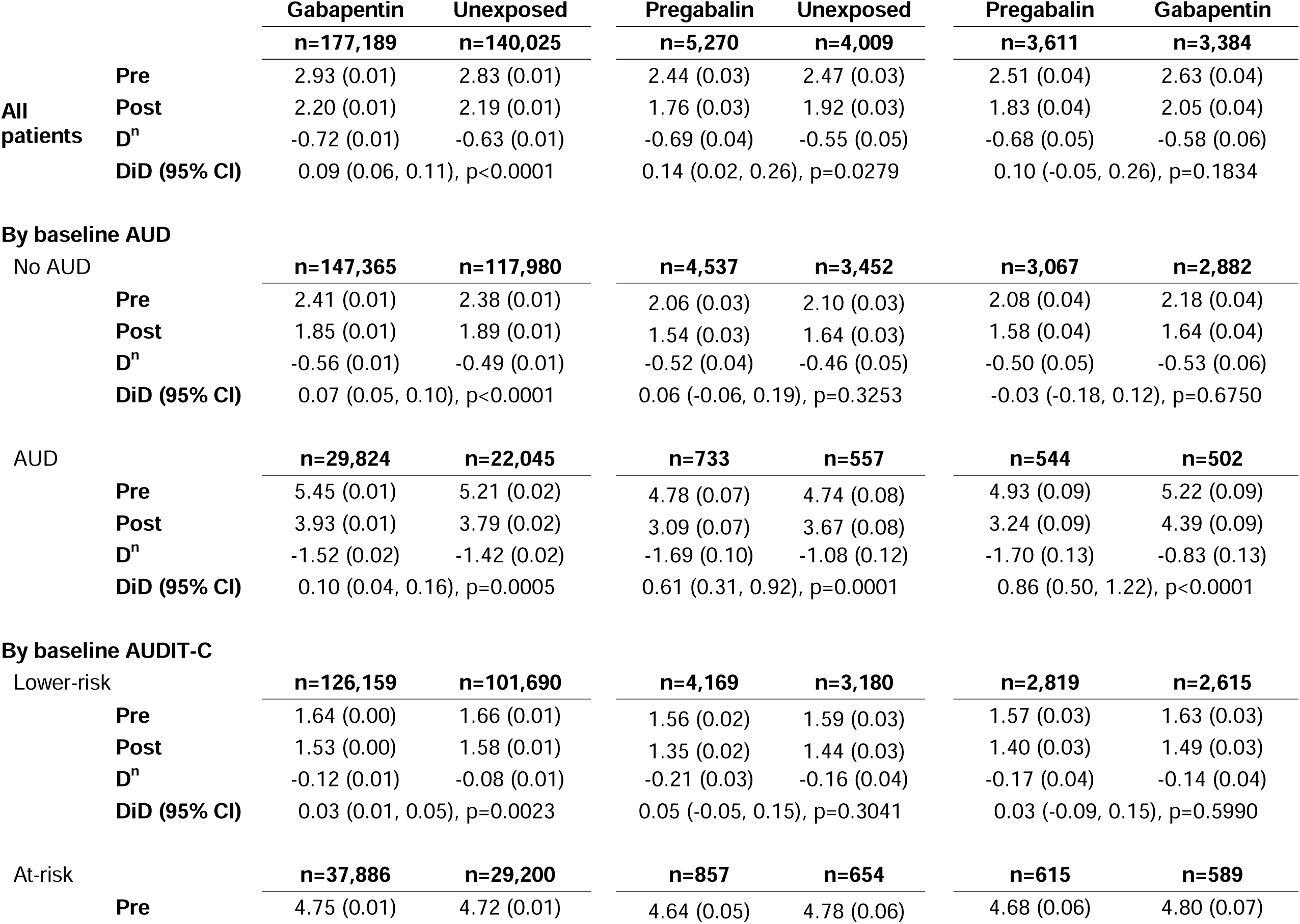
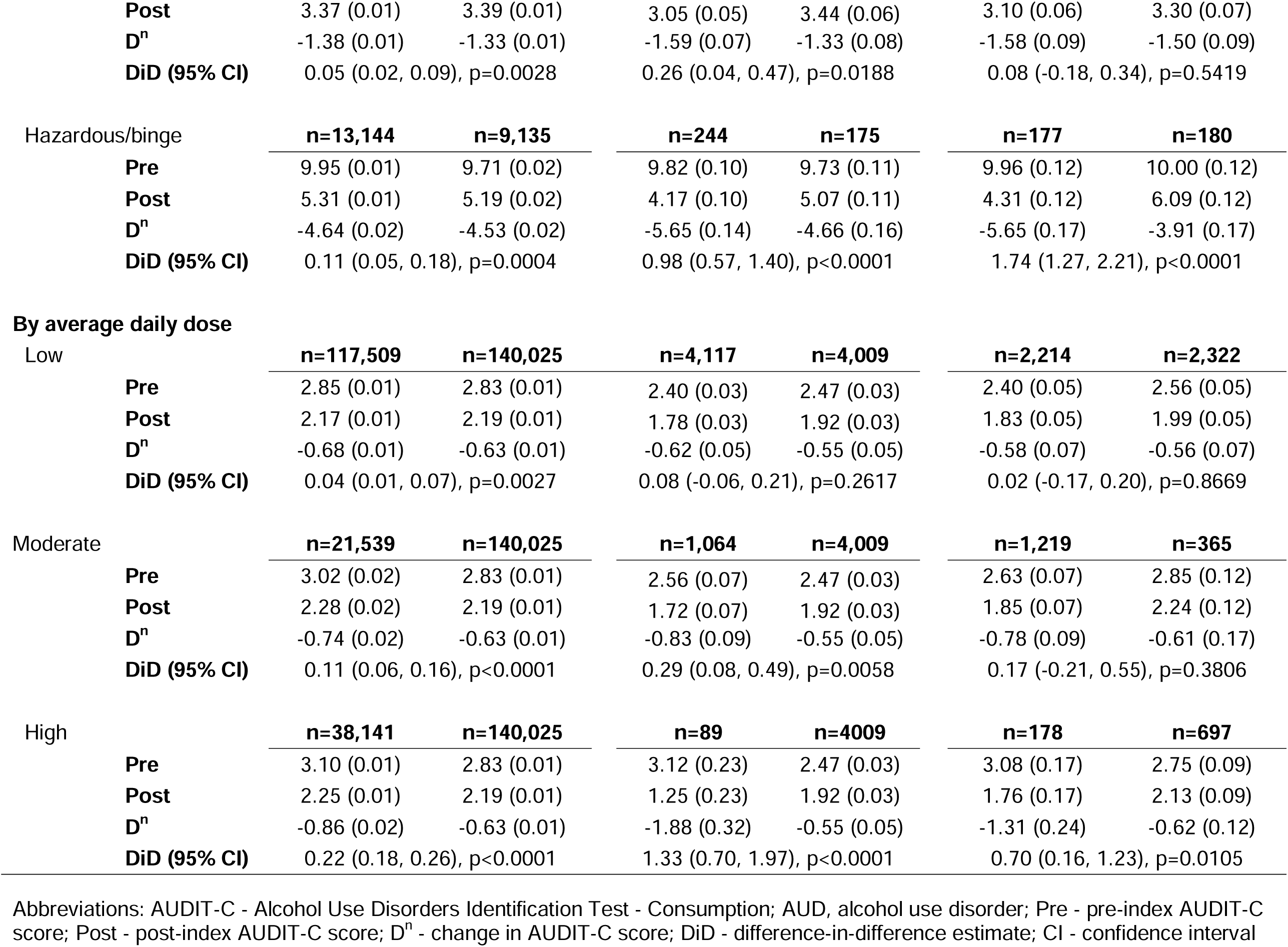
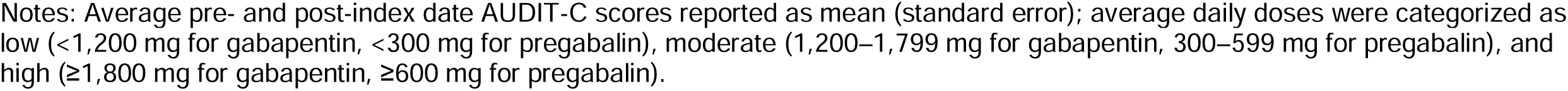
Estimated average pre- and post-index date AUDIT-C scores and difference-in-differences (DiD), overall, by current alcohol use diagnosis, and level of alcohol consumption.

|  |  | Gabapentin | Unexposed | Pregabalin | Unexposed | Pregabalin | Gabapentin |
| --- | --- | --- | --- | --- | --- | --- | --- |
|  |  | n=177,189 | n=140,025 | n=5,270 | n=4,009 | n=3,611 | n=3,384 |
| All patients | Pre | 2.93 (0.01) | 2.83 (0.01) | 2.44 (0.03) | 2.47 (0.03) | 2.51 (0.04) | 2.63 (0.04) |
|  | Post | 2.20 (0.01) | 2.19 (0.01) | 1.76 (0.03) | 1.92 (0.03) | 1.83 (0.04) | 2.05 (0.04) |
|  | D<sup>n</sup> | -0.72 (0.01) | -0.63 (0.01) | -0.69 (0.04) | -0.55 (0.05) | -0.68 (0.05) | -0.58 (0.06) |
|  | DiD (95% CI) | 0.09 (0.06, 0.11), p<0.0001 |  | 0.14 (0.02, 0.26), p=0.0279 |  | 0.10 (-0.05, 0.26), p=0.1834 |  |
| By baseline AUD |  |  |  |  |  |  |  |
| No AUD |  | n=147,365 | n=117,980 | n=4,537 | n=3,452 | n=3,067 | n=2,882 |
|  | Pre | 2.41 (0.01) | 2.38 (0.01) | 2.06 (0.03) | 2.10 (0.03) | 2.08 (0.04) | 2.18 (0.04) |
|  | Post | 1.85 (0.01) | 1.89 (0.01) | 1.54 (0.03) | 1.64 (0.03) | 1.58 (0.04) | 1.64 (0.04) |
|  | D<sup>n</sup> | -0.56 (0.01) | -0.49 (0.01) | -0.52 (0.04) | -0.46 (0.05) | -0.50 (0.05) | -0.53 (0.06) |
|  | DiD (95% CI) | 0.07 (0.05, 0.10), p<0.0001 |  | 0.06 (-0.06, 0.19), p=0.3253 |  | -0.03 (-0.18, 0.12), p=0.6750 |  |
| AUD |  |  |  |  |  |  |  |
|  |  | n=29,824 | n=22,045 | n=733 | n=557 | n=544 | n=502 |
|  | Pre | 5.45 (0.01) | 5.21 (0.02) | 4.78 (0.07) | 4.74 (0.08) | 4.93 (0.09) | 5.22 (0.09) |
|  | Post | 3.93 (0.01) | 3.79 (0.02) | 3.09 (0.07) | 3.67 (0.08) | 3.24 (0.09) | 4.39 (0.09) |
|  | D<sup>n</sup> | -1.52 (0.02) | -1.42 (0.02) | -1.69 (0.10) | -1.08 (0.12) | -1.70 (0.13) | -0.83 (0.13) |
|  | DiD (95% CI) | 0.10 (0.04, 0.16), p=0.0005 |  | 0.61 (0.31, 0.92), p=0.0001 |  | 0.86 (0.50, 1.22), p<0.0001 |  |
| By baseline AUDIT-C |  |  |  |  |  |  |  |
| Lower-risk |  | n=126,159 | n=101,690 | n=4,169 | n=3,180 | n=2,819 | n=2,615 |
|  | Pre | 1.64 (0.00) | 1.66 (0.01) | 1.56 (0.02) | 1.59 (0.03) | 1.57 (0.03) | 1.63 (0.03) |
|  | Post | 1.53 (0.00) | 1.58 (0.01) | 1.35 (0.02) | 1.44 (0.03) | 1.40 (0.03) | 1.49 (0.03) |
|  | D<sup>n</sup> | -0.12 (0.01) | -0.08 (0.01) | -0.21 (0.03) | -0.16 (0.04) | -0.17 (0.04) | -0.14 (0.04) |
|  | DiD (95% CI) | 0.03 (0.01, 0.05), p=0.0023 |  | 0.05 (-0.05, 0.15), p=0.3041 |  | 0.03 (-0.09, 0.15), p=0.5990 |  |
| At-risk |  | n=37,886 | n=29,200 | n=857 | n=654 | n=615 | n=589 |
|  | Pre | 4.75 (0.01) | 4.72 (0.01) | 4.64 (0.05) | 4.78 (0.06) | 4.68 (0.06) | 4.80 (0.07) |
|  | **Post** | 3.37 (0.01) | 3.39 (0.01) | 3.05 (0.05) | 3.44 (0.06) | 3.10 (0.06) | 3.30 (0.07) |
|  | **D<sup>n</sup>** | -1.38 (0.01) | -1.33 (0.01) | -1.59 (0.07) | -1.33 (0.08) | -1.58 (0.09) | -1.50 (0.09) |
|  | **DiD (95% CI)** | 0.05 (0.02, 0.09), p=0.0028 |  | 0.26 (0.04, 0.47), p=0.0188 |  | 0.08 (-0.18, 0.34), p=0.5419 |  |
| Hazardous/binge |  | **n=13,144** | **n=9,135** | **n=244** | **n=175** | **n=177** | **n=180** |
|  | **Pre** | 9.95 (0.01) | 9.71 (0.02) | 9.82 (0.10) | 9.73 (0.11) | 9.96 (0.12) | 10.00 (0.12) |
|  | **Post** | 5.31 (0.01) | 5.19 (0.02) | 4.17 (0.10) | 5.07 (0.11) | 4.31 (0.12) | 6.09 (0.12) |
|  | **D<sup>n</sup>** | -4.64 (0.02) | -4.53 (0.02) | -5.65 (0.14) | -4.66 (0.16) | -5.65 (0.17) | -3.91 (0.17) |
|  | **DiD (95% CI)** | 0.11 (0.05, 0.18), p=0.0004 |  | 0.98 (0.57, 1.40), p<0.0001 |  | 1.74 (1.27, 2.21), p<0.0001 |  |
| **By average daily dose** |  |  |  |  |  |  |  |
| Low |  | **n=117,509** | **n=140,025** | **n=4,117** | **n=4,009** | **n=2,214** | **n=2,322** |
|  | **Pre** | 2.85 (0.01) | 2.83 (0.01) | 2.40 (0.03) | 2.47 (0.03) | 2.40 (0.05) | 2.56 (0.05) |
|  | **Post** | 2.17 (0.01) | 2.19 (0.01) | 1.78 (0.03) | 1.92 (0.03) | 1.83 (0.05) | 1.99 (0.05) |
|  | **D<sup>n</sup>** | -0.68 (0.01) | -0.63 (0.01) | -0.62 (0.05) | -0.55 (0.05) | -0.58 (0.07) | -0.56 (0.07) |
|  | **DiD (95% CI)** | 0.04 (0.01, 0.07), p=0.0027 |  | 0.08 (-0.06, 0.21), p=0.2617 |  | 0.02 (-0.17, 0.20), p=0.8669 |  |
| Moderate |  | **n=21,539** | **n=140,025** | **n=1,064** | **n=4,009** | **n=1,219** | **n=365** |
|  | **Pre** | 3.02 (0.02) | 2.83 (0.01) | 2.56 (0.07) | 2.47 (0.03) | 2.63 (0.07) | 2.85 (0.12) |
|  | **Post** | 2.28 (0.02) | 2.19 (0.01) | 1.72 (0.07) | 1.92 (0.03) | 1.85 (0.07) | 2.24 (0.12) |
|  | **D<sup>n</sup>** | -0.74 (0.02) | -0.63 (0.01) | -0.83 (0.09) | -0.55 (0.05) | -0.78 (0.09) | -0.61 (0.17) |
|  | **DiD (95% CI)** | 0.11 (0.06, 0.16), p<0.0001 |  | 0.29 (0.08, 0.49), p=0.0058 |  | 0.17 (-0.21, 0.55), p=0.3806 |  |
| High |  | **n=38,141** | **n=140,025** | **n=89** | **n=4009** | **n=178** | **n=697** |
|  | **Pre** | 3.10 (0.01) | 2.83 (0.01) | 3.12 (0.23) | 2.47 (0.03) | 3.08 (0.17) | 2.75 (0.09) |
|  | **Post** | 2.25 (0.01) | 2.19 (0.01) | 1.25 (0.23) | 1.92 (0.03) | 1.76 (0.17) | 2.13 (0.09) |
|  | **D<sup>n</sup>** | -0.86 (0.02) | -0.63 (0.01) | -1.88 (0.32) | -0.55 (0.05) | -1.31 (0.24) | -0.62 (0.12) |
|  | **DiD (95% CI)** | 0.22 (0.18, 0.26), p<0.0001 |  | 1.33 (0.70, 1.97), p<0.0001 |  | 0.70 (0.16, 1.23), p=0.0105 |  |

Abbreviations: AUDIT-C - Alcohol Use Disorders Identification Test - Consumption; AUD, alcohol use disorder; Pre - pre-index AUDIT-C score; Post - post-index AUDIT-C score; D<sup>n</sup> - change in AUDIT-C score; DiD - difference-in-difference estimate; CI - confidence interval
Notes: Average pre- and post-index date AUDIT-C scores reported as mean (standard error); average daily doses were categorized as low (<1,200 mg for gabapentin, <300 mg for pregabalin), moderate (1,200–1,799 mg for gabapentin, 300–599 mg for pregabalin), and high ( ≥ 1,800 mg for gabapentin, ≥ 600 mg for pregabalin).

### Changes in alcohol consumption – Contrast B (Pregabalin vs. Unexposed)

Patients from both groups reported a reduction in AUDIT-C scores (pregabalin: D^n^ = −0.69 [SE=0.04]; unexposed: D^n^ = −0.55 [SE=0.05]). Thus, patients who initiated pregabalin had a significantly greater reduction in AUDIT-C scores than unexposed comparators (DiD: 0.14 points, 95% CI: 0.02, 0.26, p = 0.0279; Table 2, Figure 2B). This effect was greater for patients with a diagnosis of AUD at baseline (DiD: 0.61 points, 95% CI: 0.31, 0.92, p=0.0001) and those who reported baseline hazardous/binge consumption (DiD: 0.98 points, 95% CI: 0.57, 1.40, p<0.0001; Table 2, Figure 2B). A dose-response relationship was observed, with no effect at low average daily doses, an effect at moderate doses (DiD: 0.29 points, 95% CI: 0.08, 0.49, p=0.0058), and the largest effect at high doses (DiD: 1.33 points, 95% CI: 0.70, 1.97, p<0.0001).

### Changes in alcohol consumption – Contrast C (Pregabalin vs. Gabapentin)

Patients from both groups reported a reduction in AUDIT-C scores (pregabalin: D^n^ = −0.68 [SE=0.05]; gabapentin: D^n^ = −0.58 [SE=0.06]). Overall, there was no statistical difference in changes in AUDIT-C scores between pregabalin initiators and gabapentin initiators (DiD: 0.10 points, 95% CI: −0.05, 0.26, p=0.1834; Table 2, Figure 2C). However, there was strong evidence that pregabalin initiators had a greater reduction in AUDIT-C scores than gabapentin initiators among those with a diagnosis of AUD at baseline (DiD: 0.86 points, 95% CI: 0.50, 1.22, p<0.0001) and those who reported baseline hazardous/binge consumption (DiD: 1.74 points, 95% CI: 1.27, 2.21, p<0.0001; Table 2, Figure 2C). Comparative effectiveness between gabapentin and pregabalin was evident at high average daily doses (DiD: 0.70 points, 95% CI: 0.16, 1.23, p=0.0105), with no observed differences at low or moderate doses.

## Discussion

We compared changes in alcohol use among patients who initiated gabapentin or pregabalin for any indication. Using real-world EHR data from the largest integrated U.S. healthcare system, we applied propensity-score matching to balance baseline characteristics across three exposure contrasts and used DiD analyses to compare changes in drinking captured by routinely-collected AUDIT-C scores. Individuals who initiated gabapentin reported, on average, lower alcohol use than unexposed comparators; this difference did not differ as a function of AUD diagnosis or AUDIT-C risk category. Individuals who initiated pregabalin also reported a greater reduction in alcohol use than unexposed comparators, with a greater effect observed among those with AUD and those reporting the highest alcohol use at baseline. In a direct comparison, pregabalin initiators reported significantly greater reductions in alcohol use than gabapentin initiators, and those with AUD and those reporting the highest alcohol use at baseline experienced the greatest decrease. Additionally, we observed a dose-dependent relationship for both medications, with greater reductions in alcohol use among patients prescribed higher doses of either gabapentin or pregabalin. These results provide evidence of the effectiveness of either gabapentin or pregabalin to reduce alcohol use, with pregabalin as more effective than gabapentin, especially among patients who may benefit the most, i.e., those with AUD or those who report hazardous or heavy episodic drinking.

Our findings build upon and extend prior work on the use of gabapentinoids for treating AUD. Gabapentin has previously been shown to reduce craving and heavy drinking among individuals with AUD, though effects were not always consistent across all clinical trials ^20,22–30^. We previously estimated the real-world effectiveness of gabapentin using similar methods in a different VA cohort enriched with people living with HIV ^60^. In the present study, we replicated our previous findings in a broader, more generalizable cohort including all patients who have accessed VA care and expanded our analysis to compare its effectiveness against pregabalin.

Similarly, prior clinical trials of pregabalin have shown inconsistent but promising results. Pregabalin was previously shown to reduce heavy drinking frequency ^36^, and to have mixed effects on alcohol withdrawal and craving ^35,37–40^. Our study extends prior work by leveraging a large, real-world EHR dataset and applying statistical approaches that minimize confounds and approximate causal effects. We found evidence for the use of pregabalin to reduce alcohol use especially in patients with AUD. Additionally, this was the first study to directly compare the effects of gabapentin to pregabalin on reducing alcohol consumption, which addresses an important gap in the literature regarding the relative effectiveness of these two medications.

Machine learning-based secondary analyses of the Falk et al.’s multisite RCT ^29^ have suggested potential baseline factors that could help identifying treatment responders among those people with AUD receiving gabapentin ^61,62^. Additional studies are needed to shed light on potential subgroup(s) of patients who may respond to gabapentin and/or pregabalin, such as those based on genetic liability for alcohol consumption. For example, it has been shown that patients with history of alcohol withdrawal symptoms respond best to gabapentin ^23^ and preliminary works suggests that gabapentin may help people with other substance use disorders such as cannabis user disorder ^63^. Additionally, gabapentin may provide additional benefits on protracted withdrawal, alcohol-related sleep disturbances and negative affect ^64,65^. Furthermore, these medications may offer an alternative for patients with AUD and comorbid neuropathic pain, or seizure conditions, provided that use is closely monitored and controlled.

The use of both gabapentin and pregabalin use carries some risks. Pregabalin has a higher recognized abuse liability consistent with it being a Schedule V controlled drug in the United States, while gabapentin, though federally unscheduled, has been associated with misuse in certain populations (e.g., in those with AUD and comorbid opioid use disorder; ^66^) and is regulated in some states ^42,67^. In addition, our previous work leveraging the same data source as the present study found that gabapentin use was associated with increased risk of falls, fractures, and altered mental status ^68^. These established safety profiles underscore the need for careful monitoring and judicious prescribing for both medications. Importantly, our findings suggest modulation of the α2δ subunit may be a promising therapeutic pathway. The side-effect and misuse risks of current gabapentinoids highlight the potential value of developing or identifying alternative agents targeting similar mechanisms with improved safety profiles.

Although the magnitude of effect for gabapentin was comparable to that observed in other repurposing studies for AUD, including recent work that evaluated glucagon-like peptide-1 receptor agonists ^69^ and spironolactone ^70^, the effects observed for pregabalin, particularly among individuals with AUD and hazardous drinking, were larger than those typically reported in the literature. These findings suggest that while gabapentin’s effects may be modest at the population level, targeting the α2δ calcium channels may hold greater promise, either through more potent agents or optimized treatment strategies. Future studies could investigate whether the effect of gabapentinoids may become more robust if these medications are combined with others; for example, an initial clinical study suggested so when combining gabapentin with naltrexone ^71^. Similar approaches could be taken with other emerging new targets under investigation in AUD ^72,73^.

This study’s strengths include very large sample sizes and that its results are generalizable to real-world patients with different comorbid medical and mental health conditions and taking concurrent medications. However, there are limitations to the study. First, despite our use of propensity-score matching to control for baseline differences between groups, we cannot rule out unmeasured variables that could confound the results, such as patient motivation to reduce alcohol use and participation in non-pharmacological treatments. Second, this study was conducted on Veterans receiving VA care, who are, on average, older and have a higher prevalence of chronic health conditions and risk behaviors than the general US population ^74–76^. However, previous research has established that after adjusting for age, sex, race, ethnicity, region, and residence type, all of which were accounted for in this study, total disease burden between Veterans and non-Veterans does not differ ^76^. Thus, effects reported in this study may be generalizable to non-Veteran populations. Third, although individuals in VA care represent a diversity of backgrounds, women represented a small proportion of individuals in the cohort, which prevented us from examining sex differences. Finally, gabapentin and pregabalin were examined without restriction to indication. The inability to capture the clinical rationale for prescribing limited adjustment for indication-related factors, which may have introduced residual confounding by indication in contrasts with unexposed comparators.

In conclusion, we found that both gabapentin and pregabalin are associated with reductions in drinking in a real-world setting, with stronger evidence for the effectiveness of pregabalin particularly for patients with diagnosed AUD or those who report hazardous or heavy episodic drinking. Randomized controlled trials directly comparing gabapentin to pregabalin are necessary to confirm these findings.

## Data Availability

All data produced in the present study are available upon reasonable request to the authors.

## Disclaimer

The contents of this publication are the sole responsibility of the authors and do not necessarily reflect the official policy or position of the Uniformed Services University of the Health Sciences, the Department of Defense, the U.S. Department of Veterans Affairs, or Henry M. Jackson Foundation for the Advancement of Military Medicine.

## Acknowledgements

This work uses data provided by patients and collected by the Veterans Affairs (VA) as part of their care and support.

