## Supplemental Figure 1 for "Comparative effectiveness of gabapentin and pregabalin on reduction in alcohol use: A nationwide observational cohort study"

**Figure S1.** Propensity score distributions before and after matching for gabapentin vs unexposed, pregabalin vs unexposed contrasts, and gabapentin vs pregabalin.

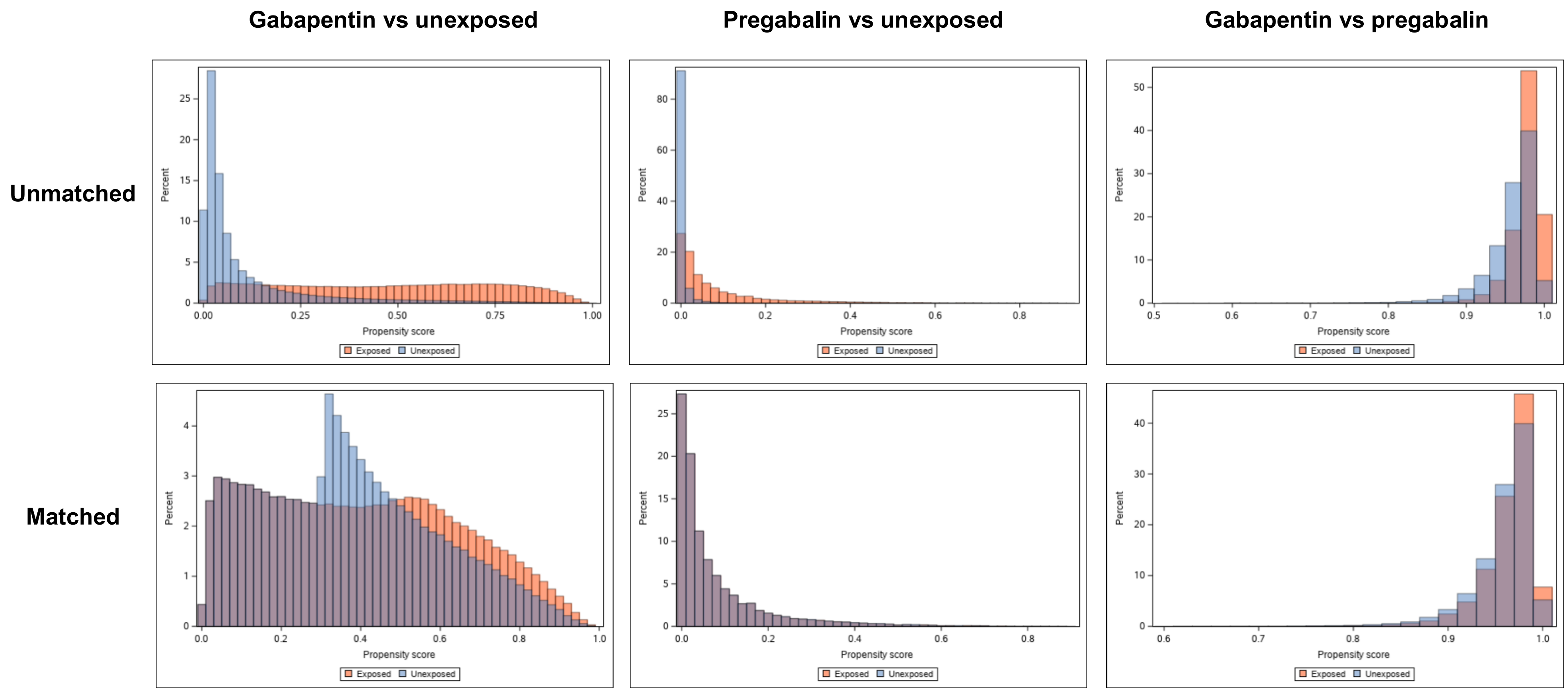
